# Integrating lifestyle, vascular, brain and cognitive markers: an exploratory multimodal approach to brain aging

**DOI:** 10.64898/2026.01.08.26343661

**Authors:** Mark R. van Loenen, Lianne B. Remie, Mara P.H. van Trijp, Mechteld M. Grootte Bromhaar, José P. Marques, Jurgen A.H.R. Claassen, Wilma T. Steegenga, Joukje M. Oosterman, Esther Aarts

## Abstract

**Background:** Lifestyle modifiable risk factors are promising intervention targets to delay cognitive decline and dementia in older adults. While previous studies have reported associations between lifestyle, vascular health, (MRI-based) brain health and cognition, few have considered all domains simultaneously. In this exploratory cross-sectional study, we examined pathways linking lifestyle to cognition via vascular and brain health, and identified key variables contributing to shared variance between these domains.

**Methods:** We included 176 Dutch adults, aged 60–75 years old with modifiable risk of cognitive decline. Composite domain scores for lifestyle, vascular health, (MRI-based) brain health, and cognitive functioning were calculated. We used Structural Equation Modelling (SEM) to test direct and indirect pathways between composite domain scores, Canonical Correlation Analysis (CCA) to identify individual variables driving shared variance between domains, and Pearson’s correlations to explore pairwise associations.

**Results:** SEM showed associations between lifestyle and vascular health (β = 0.23, *p* = 0.002), and vascular and brain health (β = 0.18, *p* = 0.01). No direct pathways were found between lifestyle and brain health, or between vascular health and cognitive functioning. CCA revealed vascular markers, specifically BMI (-0.837), blood pressure (-0.837), and blood lipids (-0.408 to -0.466), as the strongest drivers of shared variance with cerebral perfusion (-0.318 to -0.364).

**Discussion:** Our results indicate stepwise links between lifestyle, vascular health, and brain health. Integrating multimodal markers of all domains highlights the key role of vascular health in understanding lifestyle-related effects on brain health.

**HIGHLIGHTS:** · Brain health links to lifestyle via vascular health markers

· Lifestyle or vascular health alone is not associated with cognitive functioning

· BMI, blood pressure and blood lipids most strongly linked to cerebral blood flow

· Multimodal designs enable analysis of multifactorial causes of cognitive decline

**CLINICAL TRIAL REGISTRATION:** HELI study ClinicalTrials.gov ID: NCT05777863. Medical Ethics Research Committee (MREC) Oost-Nederland ToetsingOnline filenumber: NL78263.091.21.

COMBI study Clinicaltrials.gov ID: NCT05675007. MREC Oost-Nederland ToetsingOnline filenumber: NL80063.091.22.

## 1. INTRODUCTION

The worldwide aging population causes a rapid growth in the prevalence of age-related cognitive decline and dementia. The World Health Organization (WHO) underlines the importance of targeting modifiable risk factors to reduce the risk of cognitive decline and dementia [1]. Risk factors for cognitive decline are multifaceted, arising from a complex interplay between environmental, genetic, and lifestyle-related factors [2]. Lifestyle-related risk factors, such as poor diet, physical inactivity, stress, and disturbed sleep, have been identified as promising, modifiable targets for preventive interventions.

Unhealthy lifestyle patterns contribute to cardiovascular risk factors including obesity, hypertension, hypercholesterolemia and type-II diabetes [3], and evidence shows that adopting a healthier diet and increasing physical activity can help reduce these risks [4]. Cardiovascular disease risk, in turn, is a well-established predictor of increased age-related cognitive decline and dementia [5], likely due to its effects on various distinct brain health markers, such as brain activity, perfusion and volume [6, 7]. Risk factors of cardiovascular disease negatively affect specific higher-order cognitive functions, such as working memory and episodic memory. They also impact the structure and function of associated brain regions, such as the hippocampus and dorsolateral prefrontal cortex (dlPFC) [6]. Together, this indicates a link between lifestyle, cardiovascular disease risk factors and cognitive outcomes, as well as an intermediate role for brain structure and function [6, 8–11].

While numerous studies have investigated bilateral associations between pairs of domains, such as lifestyle and cardiovascular health, or brain health and cognitive functioning, few have integrated all of these aspects into a comprehensive model to investigate potential pathways linking lifestyle, cardiovascular health, brain health, and cognitive functioning in populations at risk for cognitive decline. The current study aimed to address this knowledge gap by investigating an integrated model that includes multiple markers related to the domains of lifestyle, vascular health, (MRI-based) brain health, and cognitive functioning in 176 older adults aged 60–75 years old with an elevated risk of cognitive decline. We hypothesized a most probable stepwise pathway in which healthier lifestyle patterns are associated with improved vascular health, which in turn benefits brain health (measured with MRI) that ultimately relates to stronger cognitive functioning. We additionally hypothesized that more direct pathways could be involved, potentially bypassing mechanisms such as lifestyle factors directly affecting markers of brain health or cognitive functioning. In addition, we investigated which individual variables most strongly accounted for the associations observed between the different domains. These analyses provide an overview of the possible relations between outcome measures related to lifestyle, vascular health, brain health, and cognitive functioning, by integrating multiple involved pathways into a more extensive and cohesive model.

## 2. MATERIALS AND METHODS

### 2.1 Participants

For this study, baseline data was sourced from two studies, the HELI (n = 102) and COMBI (n = 74) studies, which are part of the MOCIA research program [12] and recruited according to similar inclusion and exclusion criteria (explained in detail previously ([13]; in press). Participants were aged between 60–75 years at moment of study inclusion. Inclusion criteria were based on the presence of at least two lifestyle-modifiable factors of the Cardiovascular Risk Factors, Aging, and Incidence of Dementia (CAIDE [14]) score: (1) body mass index (BMI) ≥25, (2) physical inactivity, (3) hypertension, (4) hypercholesterolemia, (5) diabetes type-II, and (6) mild cardiovascular disease. Exclusion criteria included history of cerebrovascular events, neurological disease, current malignant disease, and current psychiatric disorders. To exclude participants with possible undiagnosed (mild) cognitive impairment, a Telephone Interview for Cognitive Status (TICS-M1 [15]) was performed during prescreening with a cutoff score of ≥23 (of a total of 39) as an additional requirement for inclusion. Data acquisition took place between May 2022 and March 2024 at the Donders Center for Cognitive Neuroimaging of the Radboud University (RU-DCCN; Nijmegen, The Netherlands) and at the Division of Human Nutrition and Health of Wageningen University and Research (WUR-HNH; Wageningen, The Netherlands). All participants provided informed written consent.

### 2.2 Demographics

Participants completed questionnaires on demographics including age, sex, and education level. Education level was stratified in levels of low, medium and high education based on the International Standard Classification of Education (ISCED 2011 [16]).

### 2.3 Lifestyle markers

All participants completed validated questionnaires on the lifestyle domains of diet, physical activity, stress and mindfulness, and sleep.

#### 2.3.1 Diet score

A Dutch Healthy Diet (DHD) index score was calculated as a measure of adherence to the Dutch dietary guidelines 2015 [17]. The DHD-index score was derived from results of the Dutch Mediterranean-Dietary Approaches to Stop Hypertension Intervention for Neurodegenerative Delay (MIND-NL) food frequency questionnaire (FFQ) [18, 19] in HELI participants, and from a separate extensive FFQ in COMBI participants [20]. From both FFQs, a DHD-index score was calculated based on questions with regard to consumption of the following food groups: (1) vegetables, (2) fruit, (3) whole-wheat products, (4) legumes, (5) nuts, (6) dairy, (7) fish, (8) tea, (9) spreadable and cooking fats, (10) red meat, (11) processed meat, (12) sugary drinks, (13) alcohol, (14) salt, and (15) other unhealthy food choices (e.g. take-away). Each food group was graded from 0-10 points (based on a 2-day average intake [21]), resulting in a maximum of 150 points for all food groups combined. A higher DHD-index score indicates stronger adherence to the Dutch dietary guidelines 2015 [17]. One participant failed to complete the FFQ, resulting in n=1 excluded DHD-index dataset.

#### 2.3.2 Physical activity score

We assessed physical activity by using the Short Questionnaire to Assess Health-enhancing physical activity (SQUASH), which is based on the Dutch guidelines for physical activity. Participants scored their physical activity by amount of minutes per day, and days per week of specific activities in the last months. Activities included: (1) walking to work, (2) cycling to work, (3) cycling to work with electric bike, (4) light activity at work, (5) intense activity at work, (6) light home chores, (7) intense home chores, (8) cleaning home, (9) walking in free time, (10) cycling in free time, (11) cycling in free time with electric bike, (12) gardening, and (13) sport activities. For each activity an activity score was calculated based on the amounts of minutes per day and days per week spent on the activity, multiplied by a factor of metabolic equivalent of task (MET) to correct for the physical intensity of each activity. A total score was calculated of all activities multiplied by the corresponding MET factors, with a higher SQUASH-score indicating more physical activity on a weekly basis.

In addition, we used the LASA Sedentary Behavior Questionnaire (LASA-SBQ), a self-report questionnaire specifically designed to assess sedentary behavior in older adults [22]. Participants scored the average time in minutes per day spent on 10 different sedentary behavior items during weekdays and weekends. The sedentary behavior items consisted of: (1) napping on chair or couch, (2) reading while seated or lying down, (3) listening to music while seated or lying down, (4) watching television, video or DVD, (5) performing a hobby while seated (e.g. knitting, playing music), (6) talking (in-person or on the phone) while seated, (7) sitting behind a computer, (8) performing sitting activities (e.g. writing a letter, administrative tasks), (9) commuting by car, bus or train, 10) visiting activities while seated (e.g. church, theater, cinema). A higher score indicates a higher amount of sedentary behavior.

In total, data from 6 participants were excluded from analysis of these physical activity questionnaires due to clear input errors (e.g., indicating activities on more than 7 days per week, more hours of activity per day than waking hours, etc.).

#### 2.3.3 Psychological wellbeing scores

We assessed scores of psychological wellbeing in three different components: stress, mood, and mindfulness.

We used the 10-item Perceived Stress Scale (PSS) [23] to determine perceived stress levels based on a Likert scale between 0 (‘never’) and 4 (‘very often’), with total scores indicating low perceived stress (0–13), moderate perceived stress (14–26) and high perceived stress (27–40).

We used the Hospital Anxiety and Depression Scale (HADS) [24] to assess the anxiety and depression-related complaints from the past four weeks. The HADS questionnaire consists of two subscales: an anxiety-scale and a depression-scale, each with 7 questions with multiple-choice answers based on Likert scales. We summed both subscale scores into a total score. A higher score indicates a greater frequency of anxiety- or depression-related complaints.

Because of the significant positive impact of mindfulness on stress, stress perception, reduced anxiety and depression, we have additionally used the Five Facet Mindfulness Questionnaire (FFMQ) to assess multiple components of mindfulness. The FFMQ consists of five subscales: observing, describing, acting with awareness, non-judgmental (of inner experience), and non-reactivity (to inner experience). Each subscale has between 7-8 questions with multiple-choice answers based on a Likert scale between 1 (‘never or very rarely true’) and 5 (‘very often or always true’). We summed all subscale scores into a total score. A higher total score indicates greater level of mindfulness.

#### 2.3.4 Sleep quality score

The Pittsburgh Sleep Quality Index (PSQI) [25] was used to assess sleep quality. The PSQI measures sleep quality and sleep disturbance over the past month, and consists of seven components: subjective sleep quality, sleep latency, sleep duration, sleep efficiency, sleep disturbances, use of sleeping medication, and daytime dysfunction. Each component is scored between 0–3 points with a maximum of 21 points. We summed all subscale scores into a total score. A higher score indicates worse sleep quality.

### 2.4 Cardiovascular health markers

#### 2.4.1 BMI and blood pressure

Length and weight were measured to calculate BMI (kg/m^2^). Systolic blood pressure (SBP) and diastolic blood pressure (DBP) were measured using an Omron X3 Comfort digital blood pressure monitor. Blood pressure was measured two times, and three times if the first two measurements showed a significant deviation between each measurement. Multiple blood pressure measurements were averaged, and the averaged SBP and DBP values were then used to calculate mean arterial pressure (MAP = DBP + ⅓ × (SBP – DBP) [26]).

#### 2.4.2 Blood cardio-metabolic markers

Blood tubes (Vacutainers, BD) containing sodium fluoride were used to analyze glucose. Blood tubes containing lithium heparin were used to analyze insulin, total-high-density lipoprotein (HDL) cholesterol, total- low-density lipoprotein (LDL) cholesterol, and triglycerides. Blood tubes were inverted ten times, and afterwards centrifuged (3000 × g) for 8 min at 20°C. Plasma was collected and stored at -80°C until further analysis. The blood cardio-metabolic marker analyses were performed at the Clinical Chemistry and Hematology Laboratory of Gelderse Vallei Hospital (Ede, The Netherlands) using standard clinical laboratory assays. In short, glucose was analyzed in 500 µL plasma using the enzymatic assay called glucose hexokinase 3 on an Atellica™ CH analyzer with a measuring interval of 0.2–38.9 mmol/L. Insulin was analyzed with the IMMULITE^®^ 2000 Insulin solid-phase, enzyme-labeled chemiluminescent immunometric assay. We calculated a Homeostatic Model Assessment for Insulin Resistance (HOMA-IR = (fasted insulin in µU/mL × fasted glucose in mmol/L) / 22.5) value for each participant as a measure of insulin resistance. Total HDL- and LDL-cholesterol, and triglycerides, were analyzed in 500 µL plasma using the Attelica^™^ CH analyzer with a measuring interval of 0.52–3.34 mmol/L for HDL-cholesterol, 0.13–25.90 mmol/L for LDL-cholesterol, and 0.11–6.22 mmol/L for triglycerides. Blood drawing was performed in the morning, and participants were instructed to remain fasted from 20.00 PM the day before the outcome measure visit. Blood drawing was not successful for every participant, as in four cases an insufficient amount of blood could be collected for the analyses, and in one case, venipuncture was unsuccessful.

### 2.5 Brain health (neuroimaging) markers

We acquired multiple measures of brain structure and function from neuroimaging. All participants were scanned with a 3T MAGNETOM Skyra MR scanner (Siemens AG, Healthcare Sector, Erlangen, Germany) using a standard 32-channel head coil at the RU-DCCN. Total scanning time was approximately 90 minutes, including preparation and handling time. To reduce head movement during scanning, the participants’ heads were fixed using tightly fitted head cushions. A detailed overview of all sequence parameters has been provided previously ([13]; in press). We used the dorsolateral prefrontal cortex and hippocampus as specific regions of interest (ROIs).

#### 2.5.1 Structural brain imaging, parcellation and volumetrics

We acquired structural images using a T1-weighted MP2RAGE sequence (TR = 6000 ms, TE = 2.34 ms, TI1/2 = 700/2400 ms, flip angle = 6◦, voxel size = 1.0 mm isotropic). The uniform, second inversion (UNI x INV2 [27]) MP2RAGE images were processed by performing cortical reconstruction and volumetric segmentation using the ‘aparc+aseg’ segmentation pipeline from the Freesurfer image analysis suite, which is documented and freely available for download online (http://surfer.nmr.mgh.harvard.edu/).

Hippocampal volumes were extracted from the parcellated Aseg Atlas (‘aseg_stats’), and the rostral middle frontal volumes were extracted from the Desikan-Kiliany atlas (‘aparc_volume’) as a proxy for dorsolateral prefrontal cortex volume. All extracted brain volume measures are in mm^3^ and shown as a ratio of total intracranial volume (ICV).

#### 2.5.2 fMRI brain activity

We performed task-based functional magnetic resonance imaging (fMRI) to assess working memory (WM)-related neural recruitment and performance. We acquired fMRI images with a blood-oxygen-level-dependent (BOLD) contrast using a whole-brain T2*-weighted gradient-echo single-echo echo planar imaging (EPI) sequence (TR = 1500 ms, TE = 33.40 ms, flip angle = 75◦, voxel size = 2.0mm isotropic), incorporating multiband acceleration with a factor of 4. To assess WM-related neural recruitment and performance, we conducted an N-back task during fMRI [28]. During the N-back task, participants were asked to actively monitor and remember a series of numerical visual stimuli (numbers from 1 – 9) during a specific timeframe, and to give a response once the presented stimulus was identical to the stimulus presented an ‘n’ number (0, 1, or 2) of trials before. We performed a 15-minute, three-condition block design with the following conditions: 0-back (control condition; respond when stimulus showed the number 1), 1-back (respond when shown stimulus is identical to stimulus shown one number back), and 2-back (respond when shown stimulus is identical to stimulus shown two numbers back). Participants received the task instructions outside of the scanner and were able to perform a practice trial whilst an involved researcher observed to verify that the participant understood the instructions.

We used fMRIPrep (23.2.0; RRID:SCR_016216 [29]) to preprocess the anatomical and fMRI data, and subsequently performed first level and ROI analyses. Realignment and coregistration of the functional scans to the native T1-weighted anatomical scan was performed before the images were susceptibility distortion-corrected and normalized to standard MNI152 space. An extensive description of all fMRIPrep preprocessing steps is provided in the appendix (Appendix A: Supplementary Material). We used Statistical Parametric Mapping 12 (SPM12; https://www.fil.ion.ucl.ac.uk/spm/software/spm12/) in MATLAB R2024a (Mathworks Inc.; https://nl.mathworks.com/products/matlab.html) to perform spatial smoothing of the preprocessed BOLD time series using a 6 mm full-width half-maximum (FWHM) Gaussian smoothing kernel. The data quality of subject-specific fMRI datasets and task performance was evaluated before including the data in the group analysis. Participants were excluded if they appeared to have misunderstood the N-back task during scanning (failed task execution), as indicated by either less than 30% target accuracy of the 0-back (control) condition and/or more than 35% false positive responses to non-targets in any of the 0-, 1- or 2-back conditions. Furthermore, we evaluated the quality of the complete fMRI dataset by examining reports and motion parameters provided by fMRIPrep. For datasets with possible questionable data quality, indicated by a framewise displacement mean >0.5mm or max >4mm), the first-level main task (2-back – 0-back) contrast (*p* < 0.05 FWE-corrected) was visually inspected for fronto-parietal activation patterns consistent with WM. Following these quality control assessments, n = 5 participants were excluded because of failed task execution, and n = 3 participants were excluded because of questionable fMRI data quality. Additionally, n = 5 participants had no available fMRI data due to issues with the MRI-scanner during acquisition, leading to a total of n = 13 excluded fMRI datasets.

For the first-level analyses, we performed a general linear model (GLM) to model the block-related BOLD responses. The model included three condition-specific regressors based on the three N-back task conditions (0-back, 1-back, 2-back). In addition to these condition-specific regressors, we also included a number of confound regressors: framewise displacement, six realignment parameters, six anatomical principal component noise regressors (aCompCor), six temporal principal component noise regressors (tCompCor), all cosine regressors, and all independent components labeled as noise by ICA-AROMA. ICA-AROMA components were taken from fMRIPrep version 23.0.2 (RRID:SCR_002502) and incorporated into the model, as opposed to using AROMA-denoised data. To avoid removing task-related variance shared with the noise regressors, ICA-AROMA components showing >0.05 covariation with the task design were excluded from the model. Each experimental condition in the block design was modeled as a separate regressor by applying the hemodynamic response function provided by SPM. To account for low-frequency drift, cosine regressors were included instead of applying a high-pass filter. The primary WM contrast was defined as 2-back minus 0-back, and first level statistical parametric contrast maps were generated for each individual participant.

To analyse neural recruitment in the dlPFC and hippocampus, we created independent ROI masks of these brain regions. The dlPFC mask was created based on the dlPFC-specific regions associated with WM activation from a large-scale 1091-study meta-analysis (Neurosynth; https://neurosynth.org/analyses/terms/working%20memory/), which were overlaid with peak dlPFC voxels associated with aging-related N-back effects from a 96-study meta-analysis [30]. The hippocampus mask was taken from the Harvard-Oxford subcortical atlas [31]. The mean ROI β-values of all task-based regressors retrieved from SPM12 were taken and used for the group analyses.

#### 2.5.3 Cerebral blood flow

Arterial spin labelling allows for an absolute quantification of cerebral blood flow (CBF) without the use of an intravenous contrasting agent, but instead labels arterial blood water in the carotid arteries using an inversion pulse before the blood travels to and through the brain. We used a vessel scout sequence (TR = 47.90 ms, TE = 8.15 ms, flip angle = 9◦, voxel size = 1.2 × 1.2 × 6.0 mm) to visualize the carotid arteries of each participant and manually placed the tagging plane perpendicular to the feeding arteries according to anatomical landmarks. We subsequently performed a Hadamard-encoded multi-PLD pseudocontinuous ASL (pCASL) sequence to acquire perfusion maps and acquired an equilibrium magnetization (M_0_) calibration image for each participant. A necessary MRI scanner software update and subsequent sequence update in May 2023, required a change of our pCASL sequence. From March 2022 to May 2023, we used a pCASL sequence (TR = 4000 ms, TE = 16.28 ms, excitation flip angle = 120◦, refocusing flip angle = 90◦, voxel size = 1.8 × 1.8 × 3.5 mm, sub-bolus duration = 400 ms, PLD = 100 ms, repeats = 2) with water suppression enhanced through T1 effects (WET) background suppression, accompanied by an M_0_ calibration protocol (TR = 500/1000/1500ms ms, TE = 16.28 ms, flip angle = 120◦, voxel size = 1.8 × 1.8 × 3.5 mm) to be used as reference. From May 2023 to March 2024, we used the pCASL and M_0_ sequences with the same contrast parameters and total acquisition time as the original sequence but with additional undersampling during the 3D-GRASE readout present in both pCASL and M0 calibration protocols (originally 2-fold and later 2x2 with CAIPI displacement) to reduce sensitivity to subject motion. Both sequences were customer developed by the Fraunhofer Institute for Digital Medicine MEVIS, with the first being implemented in the Siemens IDEA proprietary environment and the latter being implemented in the vendor-agnostic gammaSTAR. We used the FSL BASIL ‘asl_gui’ toolset [32] to process the acquired perfusion maps and calibration images and create quantified CBF images. Perfusion images were Hadamard-decoded and motion corrected, and an ASL-mask was created to only capture voxels containing ASL signals. Using BASIL, we analyzed the 14 volumes (7 PLDs × 2 repeats) of each pre-subtracted perfusion image grouped by repeats, with a constant sub-bolus duration of 0.4 seconds (0.5 – 2.9 s). To add anatomical reference and registration to the quantified perfusion output in native space, we used the FSL_ANAT pipeline on the T1-weighted MP2RAGE structural image and supplied BASIL with the ‘T1.anat’ output directory. For calibration, we specified the M_0_ image as a proton-density calibration image (sequence TR = 1000, calibration gain = 10). The perfusion maps of all participants were visually inspected to assess data quality, and outliers of quantified perfusion were cross-checked with perfusion map quality assessment to rule out possible acquisition errors or excessive motion artifacts. Following these quality control assessments, n = 1 participant had an acquisition error (little to no signal), and n = 4 participants had excessive motion artifacts. Additionally, n = 10 participants had no available ASL data due to issues with the MRI-scanner during acquisition, leading to a total of n = 15 excluded ASL datasets.

To measure CBF in whole-brain grey matter (GM) and white matter (WM) and our ROIs of the dlPFC and hippocampus, we extracted and averaged the quantified CBF values of each voxel overlaid with a bilateral Freesurfer ASEG structural parcellation-derived GM/WM and hippocampus mask, and with the (native-space transformed) dlPFC-mask that was created for the fMRI analysis.

#### 2.5.4 Neuroinflammation

Brain myo-inositol is used as a quantifiable measure of activated microglia, used as a neuroimaging spectroscopy marker of local increased neuroinflammation [33]. We used the total quantified myo-inositol, with total water as a reference, within a voxel placed in the left dlPFC. To locally quantify mI within this voxel, we used Point RESolved Spectroscopy (PRESS) ^1^H-MRS sequence (TR = 2000 ms, TE = 35 ms, flip angle = 90◦, voxel size = 20 mm isotropic) together with chemically selective water suppression (CHESS) to suppress water proton signals. We were unable to acquire hippocampal MRS data due to insufficient spectrum quality, and thus only MRS data from the voxel within the left dlPFC was used in analysis. The voxel was manually placed in our ROIs by using a T1-weighted image to navigate. During voxel placement, we prevented the inclusion of non-brain regions within the voxel, such as cerebrospinal fluid (CSF) or the skull, as much as possible. Prior to acquisition, we performed a shimming step within the selected voxel to reduce magnetic field inhomogeneities. During shimming, we used a FWHM value of <30 as a measure of shimming success according to the Siemens MRS guidelines [34]. If time allowed, we ran the shimming procedure two times and used the lowest FWHM measurement as an indicative of spectrum quality to decide the final MRS acquisition sequence used for further preprocessing and analysis.

Output from the unsuppressed and suppressed MRS sequences were pre-processed using Osprey 2.4.0 [35], an open-source software package for pre-processing, including linear-combination modelling, tissue correction and ultimately spectrum analysis and metabolite quantification. For each participant, individual metabolite transients were frequency-corrected using lineshape reference and tissue-corrected with anatomical MP2RAGE data. The metabolite transients were then averaged to create the spectrum for further analysis. When required, eddy-current correction [36] was performed using water-unsuppressed lineshape reference data. The processed spectra were subsequently analyzed using Osprey’s linear combination modeling with default parameters. The metabolite spectra were fitted over a 0.5–4.0 ppm frequency range with 0.4 ppm knot spacing. Water reference data was separately fitted over a range of 2.0–7.4 ppm. The spectroscopy data was coregistered to the native-space T1-weighted anatomical scan for each participant, and tissue segmentation was performed using SPM12 to estimate fractional tissue volumes within the region of interest. Metabolite concentrations were then corrected for GM/WM/CSF tissue composition and relaxation times and scaled to water concentration, which ultimately generated molal concentration estimates for myo-inositol. The quantified myo-inositol values were corrected for the relative residual amplitude (relResA), a measure of spectrum quality. Manual inspections of the pre-aligned, post-aligned, averaged, and fitted spectra were performed to ensure sufficient data quality. Additionally, creatine signal-to-noise (SNR), FWHM and relResA were considered as data quality metrics. Based on the manual inspection of the spectra, and of the data quality metrics, a total of n = 7 MRS datasets were excluded.

### 2.6 Cognitive functioning markers

We administered a neuropsychological test battery incorporating a number of tests to assess the cognitive domains that are predominantly affected in cognitive aging, namely executive function, processing speed, episodic memory, and working memory. All cognitive functioning markers were corrected for education level.

#### 2.6.1 Executive function tests

The Trail Making Test (TMT) and Verbal Fluency Test (VFT) were used as measures of executive function. For the TMT, participants were provided a worksheet and were asked to connect encircled numbers by drawing a line, using a pencil. The TMT consisted of two parts: TMT-A and TMT-B. In TMT-A, participants were asked to draw a line between encircled numbers, from 1–25, in ascending order (i.e., 1 – 2 – 3 – 4 – etc.) as fast as possible. In TMT-B, participants were asked to draw a line between numbers 1–13 as fast as possible, but this time each number alternated with letters from the English alphabet in ascending order (i.e., 1 – A – 2 – B – 3 – C – etc.). For both TMT-A and TMT-B completion time were measured, and we calculated a TMT-B/TMT-A ratio score to account for TMT-A completion time and thereby obtain a more pure measure of set-shifting.

For the VFT, participants were asked to name as many different animals as possible within 60 seconds. The VFT was scored as the number of correct uniquely named animals within 60 seconds.

#### 2.6.2 Processing speed tests

The Digit Symbol Substitution Test (DSST) was used as a measure of processing speed. For the DSST, participants were provided a worksheet containing nine different symbols, each corresponding to a number between 1–9. On the same worksheet, rows of numbers between 1–9 were provided. The participants were asked to go through the numbers one-by-one, and match as many symbols to their corresponding number within 90 seconds. The DSST was scored as the amount of correctly entered symbols within 90 seconds.

#### 2.6.3 Working memory tests

The Digit Span Test (DST) and N-back task performance were used as measures of working memory. For the DST, participants were verbally presented with sequences of digits and were instructed to recall the sequence. The DST consisted of two parts: DST-forward and DST-backward. For the DST-forward, participants were asked to repeat the presented digit sequence in the exact same order. For the DST-backward, participants were asked to repeat the presented digit sequence in reverse order. The digit sequences consisted of spans of increasing lengths, starting at a digit span of two. Each span consisted of two distinct digit sequences. The maximum span length was 8 for both versions of the DST, and span length was only increased if one or two of the digit sequences in a specific span were correctly recalled. The DST was scored as the number of correctly recalled digit sequences, and we calculated a sum-score of the forward and backward DST scores.

Performance on the N-back task was assessed using d-prime (d’), calculated as *d’ = z(H) – z(F)*, where z(H) represents the z–score of the hit rate and z(F) the z-score of the false alarm rate. This signal detection theory measure considers both hit rate and false alarm rate. We calculated a contrast d’-score by subtracting the 0-back d’-score from the 2-back d’-score to measure WM-specific performance.

#### 2.6.4 Episodic memory test

The Rey Auditory Verbal Learning Test (RAVLT) was used as a measure of episodic memory. For the RAVLT, participants were verbally presented with a list of 15 different words and asked to recall as many words as possible immediately after the presentation (immediate recall). This process was repeated four times, in order for the participant to learn the different words (encoding phase). After 15 minutes, participants were again asked to recall as many of the 15 words as possible (delayed recall). We used the delayed recall outcome of the RAVLT, as we expect this measure to be more strongly affected by aging as opposed to the immediate recall outcome. The RAVLT delayed recall was scored as the number of correctly recalled words.

### 2.7 Statistical analysis

Statistical methods and analyses were performed in R version 4.0.2 (R Foundation for Statistical Computing, Vienna, Austria). We performed data quality control assessments to prevent data input errors (for questionnaires) and removed data points with poor data quality (e.g. data acquisition or processing errors). We performed an outlier analysis based on the interquartile range (IQR) method for all included variables. Specifically, values falling below the lower bound of 𝑄1 − 1.5 × 𝐼𝑄𝑅 or above the higher bound of 𝑄3 + 1.5 × 𝐼𝑄𝑅 ) were flagged as potential outliers. IQR-outliers were subsequently cross-checked against data quality control assessments. Data points that exceeded the lower- or upper IQR bound, and failed data quality control assessments or showed reported protocol deviations during acquisition, were considered invalid and were removed from the dataset. Before analysis, data normality and linearity were assessed using QQ- and scatterplots. All variables were standardized by scaling to z-scores prior to group analyses.

Baseline characteristics were calculated using mean and standard deviations (SD), or counts and percentages. Individual outcome measures were z-standardized and grouped into four outcome domains: (1) lifestyle, (2) vascular health, (3) brain health, and (4) cognitive functioning (see Table 1). For interpretation purposes, we transformed the dataset such that higher z-scores indicate better/healthier outcomes. We achieved this by applying a -1 multiplication to a number of z-standardized outcome variables for the domains of lifestyle (LASA-SBQ, PSS, HADS, PSQI), vascular health (BMI, MAP, HOMA-IR, blood LDL-cholesterol, blood triglycerides), brain health (myo-inositol dlPFC), and cognitive functioning (TMT-B/TMT-A ratio score). Composite domain scores were calculated by summing the z-scores of all individual variables. When multiple measurements assessed the same marker (e.g., several physical activity or stress-related questionnaires in the lifestyle domain), their z-scores were first averaged.

**TABLE 1.** Outcome domains and individual variables used in domain composite scores.

| **Lifestyle** | **Vascular health** | **Brain health** | **Cognitive function** |
| --- | --- | --- | --- |
| DHD-index | BMI | fMRI dIPFC | TMT |
| SQUASH | MAP | fMRI hippocampus | VFT |
| LASA-SBQ | HOMA-IR | CBF dIPFC | DSST |
| PSS | HDL-cholesterol | CBF hippocampus | DST |
| FFMQ | LDL-cholesterol | Volume dIPFC | N-back performance |
| HADS | Triglycerides | Volume hippocampus | RAVLT delayed recall |
| PSQI |  | Myo-inositol dIPFC |  |

*Abbreviations:* BMI = body mass index; CBF = cerebral blood flow; DHD = Dutch Healthy Diet; dIPFC = dorsolateral prefrontal cortex; FFMQ = Five Facet Mindfulness Questionnaire; fMRI = functional magnetic resonance imaging; HADS = Hospital Anxiety and Depression Scale; HDL = high-density lipoprotein; HOMA-IR = homeostatic model assessment of insulin resistance; LDL = low-density lipoprotein; MAP = mean arterial pressure; PSQI = Pittsburgh Sleep Quality Index; PSS = Perceived Stress Scale; LASA-SBQ = LASA Sedentary Behavior Questionnaire; SQUASH = Short Questionnaire to Assess Health-enhancing physical activity

To explore predictive relationships between the four outcome domains, we first performed simple linear regression model analyses with lifestyle as a predictor of vascular health, vascular health as a predictor of brain health, and brain health as a predictor of cognitive functioning (i.e., the hypothesized pathway). We then performed structural equation modelling (SEM) to evaluate a range of models incorporating the four outcome domains to examine each potential direct and indirect pathway within one integrated model. Additionally, we performed SEM analyses including sex as a covariate to assess its effect on the relationships between the outcome domains. We used the ‘lavaan’ and ‘lavaanplot’ R-packages to analyze the SEM model pathways and model fits. Individual model fits were assessed using a Chi-squared (χ^2^) test (p > 0.05 indicating good model fit), Comparative Fit Index (CFI), and the Root Mean Square Error of Approximation (RMSEA). A higher CFI-score or lower RMSEA-score indicated a better model fit. Model fit comparison was then performed by using the Akaike Information Criterion (AIC) and Bayesian Information Criterion (BIC), with lower AIC and BIC values indicating a better model fit. The model with the lowest AIC and BIC score was highlighted as the strongest model.

To investigate which variables within each outcome domain explain the strongest covariance between domains and potentially act as primary driving forces of associations between sets within our integrated model, we performed canonical correlation analyses (CCA). We defined the CCA variable sets X and Y based on the pathways between the outcome domains of the strongest model indicated by the SEM analyses (e.g., if the SEM analysis revealed a significant association between the lifestyle and vascular health domains, we defined the lifestyle-domain as set X and the vascular health-domain as set Y within the CCA). Prior to CCA, each variable was adjusted for sex by fitting a linear regression model with sex as the predictor and using the residuals as the corrected outcomes. For each CCA model, we first computed the correlation matrices of set X and Y. The CCAs were then performed using the ‘CCA’ R-package, including all variables of set X and Y. With CCA, canonical dimensions are computed by comparing different weighted combinations (canonical coefficients) of variables in set X and Y which show the strongest paired linear correlation. Canonical dimensions are comprised of canonical variates U and V, with corresponding canonical loadings of each individual variable. The canonical loading indicates the strength of correlation of an individual variable with canonical variate U or V, with a higher loading indicating a stronger association. A Wilk’s Lambda (*λ)* test of significance was performed on the computed canonical dimensions of each CCA, and the most significant canonical dimensions were further explored.

After conducting CCAs, we performed pairwise Pearson’s correlation analyses among all individual variables to explore specific associations between variables, beyond the broader domain-level relationships. We repeated the simple linear modeling of the total domain scores and the Pearson’s correlation analysis of all individual variables separately for men and women.

The significance threshold was set at p < 0.05 for all analyses.

## 3. RESULTS

### 3.1 Baseline demographics

Baseline characteristics of the total study sample (*N* = 176) are shown in Table 2. The most common inclusion risk factors were BMI ≥25, hypercholesterolemia, hypertension, and physical inactivity. Most of the participants had a medium-to-high educational background (n = 161) (Table 2).

**TABLE 2.** Baseline characteristics

| Characteristic | | | Available<br>datasets ($n$) |
| --- | --- | --- | --- |
| ***Demographics*** |  |  |  |
| Age (years), mean (SD) | 66.08 | (4.10) | 176 |
| Sex (female), $n$ (%) | 113 | (63.07%) | 176 |
| BMI $\geq 25$, $n$ (%) | 136 | (77.27%) | 176 |
| Physical inactivity<sup>a</sup>, $n$ (%) | 95 | (53.98%) | 176 |
| Hypertension<sup>b</sup>, $n$ (%) | 98 | (55.68%) | 176 |
| Hypercholesterolemia<sup>c</sup>, $n$ (%) | 104 | (59.09%) | 176 |
| Diabetes type-2, $n$ (%) | 28 | (15.91%) | 176 |
| Mild cardiovascular disease<sup>d</sup>, $n$ (%) | 20 | (11.36%) | 176 |
| Smoking status (current smoker), $n$ (%) | 3 | (1.70%) | 176 |
| Medication use, $n$ (%) | | | |
| Antihypertensive drugs | 61 | (34.66%) | 176 |
| Diabetes medication | 18 | (10.23%) | 176 |
| Cholesterol-lowering drugs | 34 | (19.32%) | 176 |
| Educational level<sup>e</sup>, $n$ (%) | | | 176 |
| Low | 15 | (8.52%) |  |
| Medium | 64 | (36.36%) |  |
| High | 97 | (55.11%) |  |
| ***Lifestyle domain*** |  |  |  |
| DHD diet, mean (SD) | 97.78 | (18.15) | 175 |
| SQUASH physical activity, mean (SD) | 6924.08 | (3739.51) | 169 |
| LASA-SBQ sedentary behavior, mean (SD) | 325.60 | (146.09) | 176 |
| PSS stress perception, mean (SD) | 11.97 | (4.20) | 176 |
| FFMQ mindfulness, mean (SD) | 2.98 | (0.26) | 176 |
| HADS anxiety and depression, mean (SD) | 6.97 | (5.05) | 176 |
| PSQI sleep quality, mean (SD) | 5.90 | (3.48) | 175 |
| ***Vascular health domain*** |  |  |  |
| BMI ($\text{kg}/\text{m}^2$), mean (SD) | 29.44 | (5.66) | 176 |
| MAP (mmHg), mean (SD) | 107.55 | (12.86) | 175 |
| HOMA-IR, mean (SD) | 3.72 | (2.93) | 160 |
| Blood lipid profile (mmol/L), mean (SD) |  |  |  |
| HDL-cholesterol | 1.50 | (0.40) | 173 |
| LDL-cholesterol | 3.45 | (1.04) | 173 |
| Triglycerides | 1.63 | (0.67) | 173 |
| ***Brain health domain*** |  |  |  |
| WM (N-back) fMRI activation in ROIs (au), mean (SD) |  |  |  |
| dlPFC | 0.37 | (0.15) | 162 |
| Hippocampus | -0.04 | (0.07) | 162 |
| CBF in ROIs (ml/100g/min), mean (SD) |  |  |  |
| dlPFC | 40.80 | (13.49) | 163 |
| Hippocampus | 42.81 | (11.07) | 163 |
| Brain ROI volume ratio with ICV (mm<sup>3</sup>), mean (SD) |  |  |  |
| dlPFC | 1.60 x 10<sup>-2</sup> | 1.50 x 10<sup>-3</sup> | 172 |
| Hippocampus | 4.90 x 10<sup>-3</sup> | 5.00 x 10<sup>-4</sup> | 172 |
| Myo-inositol in DLPFC (mmol/L), mean (SD) | 10.09 | (1.38) | 167 |
| ***Cognitive functioning domain*** |  |  |  |
| Trail Making Test B/A ratio, mean (SD) | 2.20 | (0.63) | 176 |
| Verbal Fluency Test, mean (SD) | 25.62 | (5.62) | 176 |
| Digit Symbol Substitution Test, mean (SD) | 52.28 | (9.96) | 176 |
| Digit Span total, mean (SD) | 15.65 | (3.27) | 176 |
| N-back task performance (2b-0b d'), mean (SD) | -1.79 | (0.99) | 172 |
| RAVLT delayed recall, mean (SD) | 7.92 | (3.12) | 176 |

*Abbreviations:* ASL = Arterial Spin Labelling; *au* = arbitrary unit; *BMI* = body mass index; *DHD* = Dutch Healthy Diet; *dlPFC* = dorsolateral prefrontal cortex; *fMRI* = functional magnetic resonance imaging; *HOMA-IR* = Homeostatic Model Assessment for Insulin Resistance; *ICV* = Intracranial volume; *MAP* = mean arterial pressure; *PSQI* = Pittsburgh Sleep Quality Index; *RAVLT* = Rey Auditory Verbal Learning Test; *ROIs* = regions of interest; *LASA-SBQ* = LASA Sedentary Behavior Questionnaire; *SD* = standard deviation; *SQUASH* = Short Questionnaire to Assess Health-enhancing physical activity.
<sup>a</sup> Based on non-adherence to the WHO Guidelines on physical activity (300 or more minutes of moderate aerobic activity or 150 minutes of vigorous aerobic activity per week)
<sup>b</sup> Based on systolic blood pressure of ≥140 mmHg and diastolic blood pressure ≥90 mmHg
<sup>c</sup> Based on total cholesterol >5mmol/L or LDL-cholesterol >3mmol/L
<sup>d</sup> Based on mild cardiovascular disease not mentioned in exclusion criteria (e.g. varicose veins, arteriosclerosis)
<sup>e</sup> Based on the International Standard Classification of Education (ISCED 2011) guidelines (low, medium, high education categorization)

### 3.2 Correlational relationships between domains of lifestyle, vascular health, brain health, and cognitive functioning

Simple linear regression modelling between the four total z-sum domain scores revealed two significant relationships. The domain of lifestyle weakly predicted vascular health (*β* = 0.41, *SE* = 0.13, *p* = 0.002), and the model explained 4.7% of variance (*R^2^* = 0.047) (Figure 1). The domain of vascular health weakly predicted brain health (*β* = 0.16, *SE* = 0.06, *p* = 0.01), and the model explained 3% of variance (R^2^ = 0.03). There was no predictive value of the domain of brain health on cognitive functioning.

**FIGURE 1.**
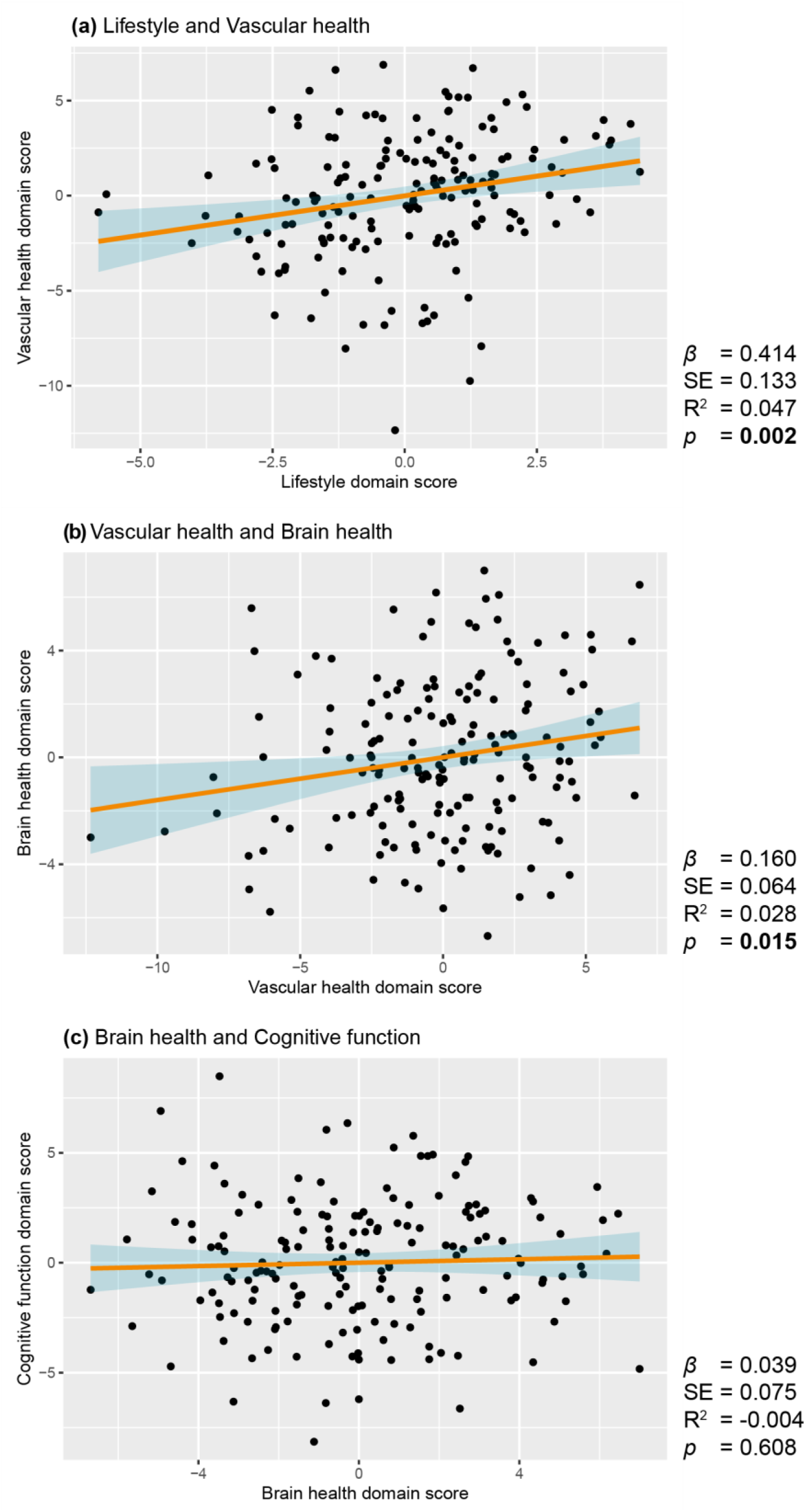
Associations between z-summed total domain composite scores. **(a-c)** Linear regression plots showing relationships between domain composite scores of (a) lifestyle and vascular health, (b) vascular health and brain health, and (c) brain health and cognitive functioning. Shaded areas indicate 95% confidence interval. The 95% confidence interval is indicated by shaded areas. R^2^ is reported as adjusted R^2^.

### 3.3 Direct and indirect pathways between lifestyle, vascular risk factors, brain health and cognitive functioning

To assess any possible indirect and direct pathways between the four total domain scores, we constructed eight different SEM models (Figure 2). Models 1 – 7 did not significantly differ from the observed data (χ^2^-test p > 0.350), indicating good model fit. Model 1 (pathway between lifestyle and cognitive functioning through vascular and brain health, without other direct pathways) showed the strongest SEM model fit based on lowest comparative model fit indices AIC and BIC (Table 3).

**FIGURE 2.**
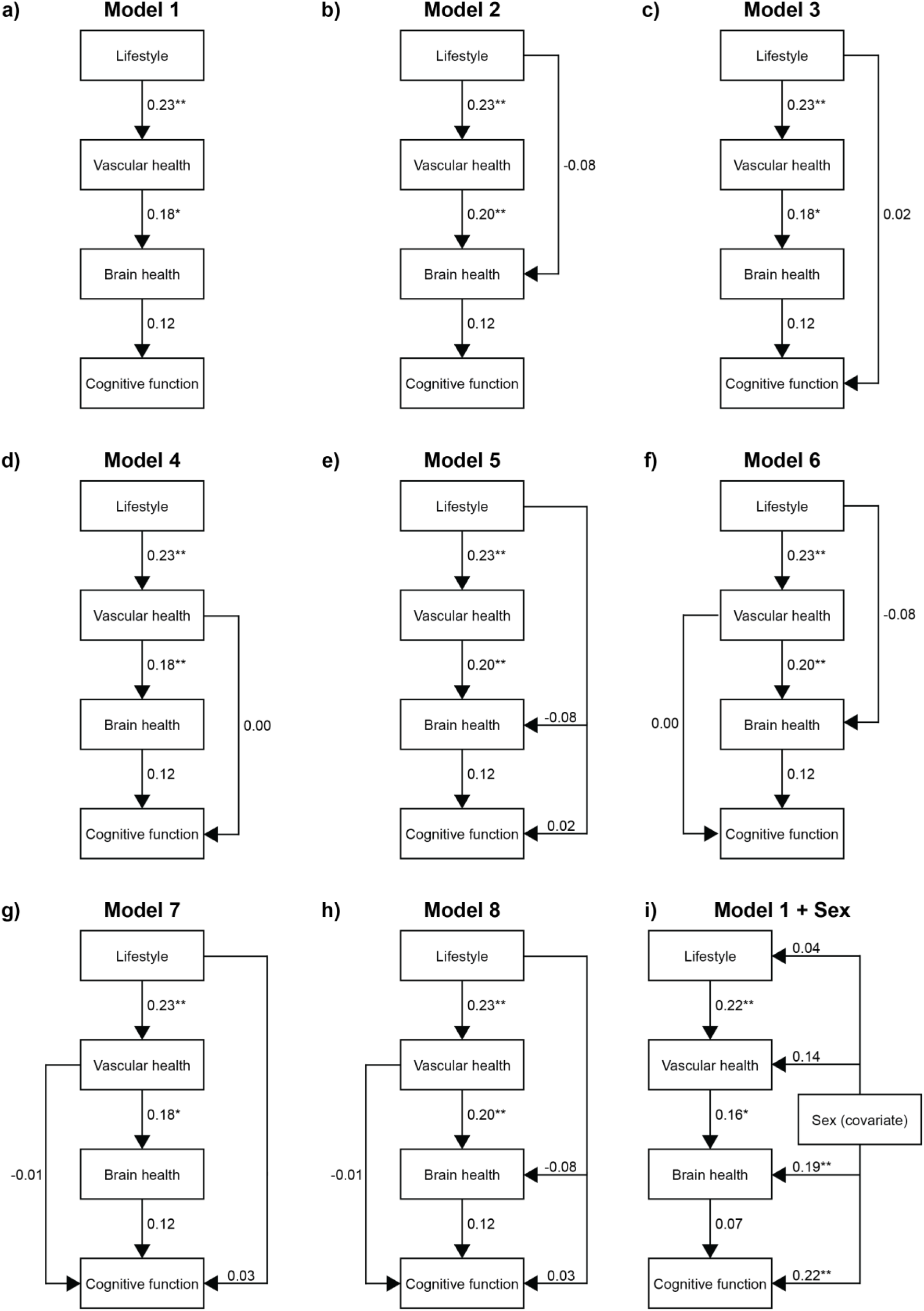
SEM models with all four domain scores. **(a-h)** SEM models containing the four domain scores with possible direct and indirect pathway combinations. **(i)** Model containing pathways of strongest fitting SEM model (Model 1), with addition of *Sex* as a covariate. * = *p*<0.05, ** = *p*<0.01

**TABLE 3.**
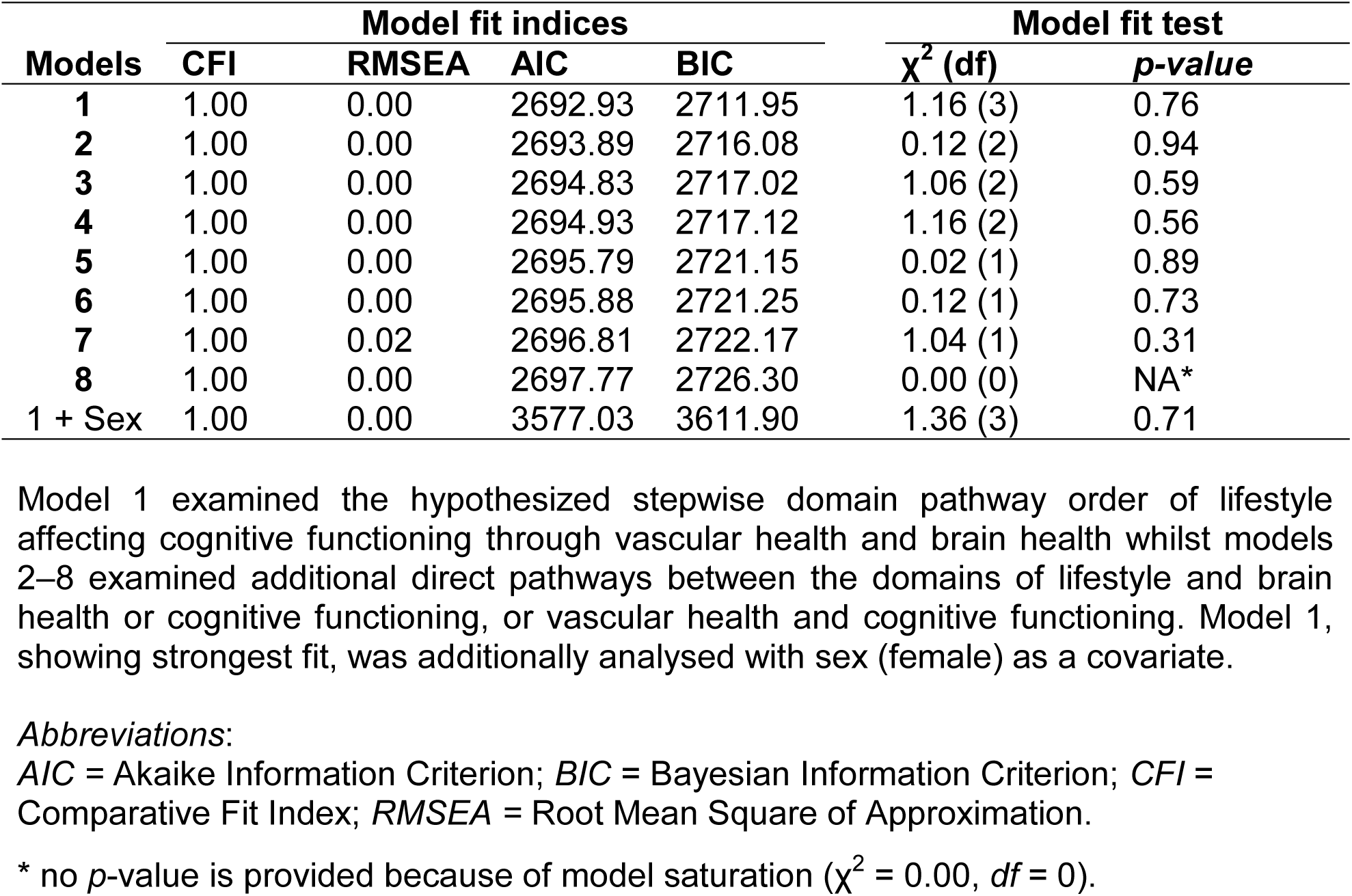
Model fit indices and model fit tests of all constructed SEM models.

In Model 1, significant shared variance was observed between the sets of lifestyle and vascular health (β = 0.23, *p* = 0.002) and between vascular health and brain health (β = 0.18, *p* = 0.01). There was no significant shared variance between the sets of brain health and cognitive functioning. Adding sex (female) as a covariate to Model 1 (Figure 2i) showed that sex significantly explains variance within the sets of brain health (β = 0.19, *p* <0.01) and cognitive functioning (β = 0.22, *p* <0.01). The AIC and BIC comparative model fit indices for this model were higher than Model 1 (without added covariate), indicating poorer model fit.

### 3.4 Shared variance between individual outcomes of lifestyle, vascular health, brain health, and cognitive functioning

To investigate which variables within each outcome domain explain the strongest covariance between domains, we constructed four different CCA models based on the associations found between the outcome sets in SEM model 1. We found no significant canonical correlations between the outcome sets of lifestyle and vascular health, or brain health and cognitive functioning (Appendix A: Supplementary Tables 1A–B, 2A–B). The CCA analysis between the domains of vascular health and brain health showed a trend-significant canonical correlation (Table 4). The first canonical dimension was trend-significant (*λ* = 0.63, *F*(42, 548) = 1.336, *p* = 0.081), indicating that around 18.6% (*r* = 0.431) of variance within the brain health set can be explained by variables from the vascular health set within this dimension. The other five dimensions showed no significant canonical correlations, and therefore only canonical dimension 1 was analyzed further.

**TABLE 4.** Tests of canonical dimensions of CCA model of vascular health and brain health domains.

| **Dimension** | **Canonical coefficient** | **Approximate *F*-statistic** | **Degrees of freedom** | **Wilk's ($\lambda$) test-statistic** | ***p*-value** |
| --- | --- | --- | --- | --- | --- |
| **1** | 0.431 | 1.336 | 42, 548 | 0.633 | 0.081 |
| **2** | 0.324 | 1.019 | 30, 470 | 0.777 | 0.441 |
| **3** | 0.273 | 0.854 | 20, 392 | 0.868 | 0.646 |
| **4** | 0.195 | 0.640 | 12, 315 | 0.938 | 0.807 |
| **5** | 0.154 | 0.501 | 6, 240 | 0.975 | 0.807 |
| **6** | 0.032 | 0.061 | 2, 121 | 0.999 | 0.941 |

*Canonical dimensions 1–6:* Linear combinations of variables (dimensions) that maximize shared variance between two variable sets. Dimension 1 captures the strongest shared variance, followed by dimension 2, and so on.

Table 5 shows the canonical loadings and standardized canonical correlation coefficients of the variables within canonical dimension 1 (for dimensions 2–5, see Appendix A: Supplementary Table 2B). Within the vascular health domain variables, MAP showed the highest canonical loading magnitude (-0.837; strong effect size), indicating that MAP (blood pressure) has the strongest association with the canonical variate of dimension 1 (U1) compared with other variables. BMI, HOMA-IR, LDL-cholesterol and triglycerides all show similar canonical loading magnitude of small-to-moderate effect sizes (-0.370 – -0.466). HDL-cholesterol shows the lowest association with canonical variate U1. For the brain health domain variables, CBF in the DLPFC and hippocampus showed the strongest canonical loading magnitude (-0.318 – -0.364; moderate effect size), indicating that the measures of local CBF (perfusion) have the strongest association with the canonical variate of dimension 1 (Y1) compared with other variables. DLPFC volume showed a small canonical loading magnitude, and WM fMRI activation in DLPFC and hippocampus and myo-inositol in DLPFC showed negligible associations with canonical variate Y1 (Table 5). Participants with a poorer vascular health status (indicated by lower MAP, BMI, HOMA-IR, LDL-cholesterol and triglycerides scores) showed worse perfusion (indicated by lower CBF scores) in the DLPFC and hippocampus, as well as a larger DLPFC volume.

**TABLE 5.**
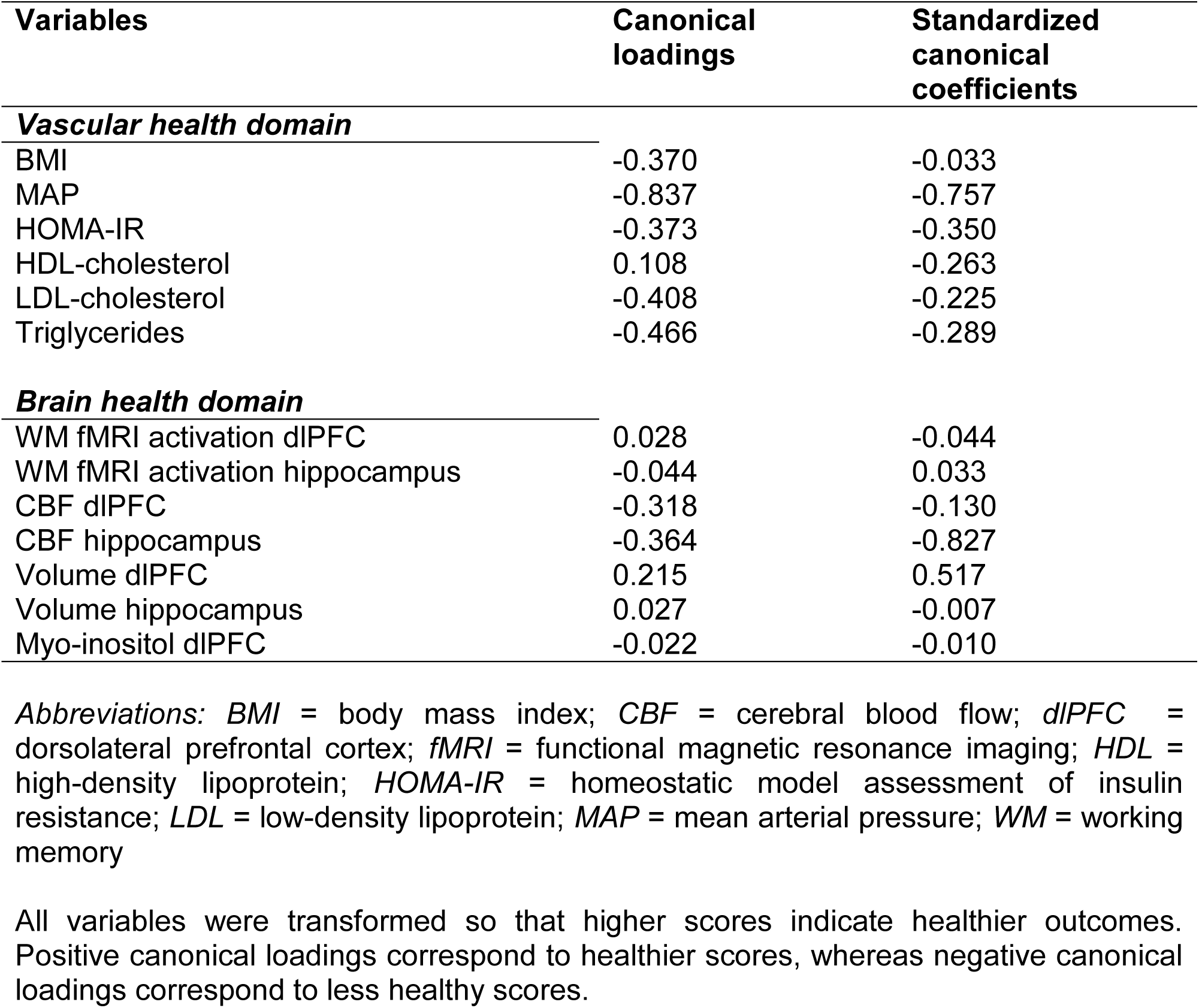
Canonical loadings and standardized canonical coefficients of dimension 1 between vascular health and brain health markers

| **Variables** | **Canonical loadings** | **Standardized canonical coefficients** |
| --- | --- | --- |
| ***Vascular health domain*** |  |  |
| BMI | -0.370 | -0.033 |
| MAP | -0.837 | -0.757 |
| HOMA-IR | -0.373 | -0.350 |
| HDL-cholesterol | 0.108 | -0.263 |
| LDL-cholesterol | -0.408 | -0.225 |
| Triglycerides | -0.466 | -0.289 |
| ***Brain health domain*** |  |  |
| WM fMRI activation dIPFC | 0.028 | -0.044 |
| WM fMRI activation hippocampus | -0.044 | 0.033 |
| CBF dIPFC | -0.318 | -0.130 |
| CBF hippocampus | -0.364 | -0.827 |
| Volume dIPFC | 0.215 | 0.517 |
| Volume hippocampus | 0.027 | -0.007 |
| Myo-inositol dIPFC | -0.022 | -0.010 |

*Abbreviations:* BMI = body mass index; CBF = cerebral blood flow; dIPFC = dorsolateral prefrontal cortex; fMRI = functional magnetic resonance imaging; HDL = high-density lipoprotein; HOMA-IR = homeostatic model assessment of insulin resistance; LDL = low-density lipoprotein; MAP = mean arterial pressure; WM = working memory
All variables were transformed so that higher scores indicate healthier outcomes. Positive canonical loadings correspond to healthier scores, whereas negative canonical loadings correspond to less healthy scores.

### 3.5 Correlational relationships between individual lifestyle, vascular health, brain health and cognitive functioning markers

To explore potential specific associations between individual variables of each domain, beyond the broader domain-level relationships, we performed a pairwise Pearson’s correlation analysis on all included variables (Figure 3). We observed a cluster of small correlations between better diet score and BMI (*r* = 0.22), HOMA-IR (*r* = 0.16), HDL-cholesterol (*r* = 0.23) and triglycerides (*r* = 0.20). These correlations indicate that lower BMI, lower insulin resistance, higher HDL-cholesterol, and lower triglycerides are associated with higher dietary score. A cluster of small associations was also observed between CBF in the dlPFC and hippocampus and vascular health domain variables. Specifically, CBF in the dlPFC was associated with BMI (*r* = 0.21), MAP (*r* = 0.26), and HOMA-IR (*r* = 0.17). Similarly, CBF in the hippocampus was associated with BMI (*r* = 0.20) and MAP (*r* = 0.30). In addition, a weak negative correlation between hippocampal CBF and physical activity score (*r* = -0.16) was observed. These correlations indicate that lower BMI, lower MAP, and lower HOMA-IR are associated with higher CBF in hippocampus and/or dlPFC, and that lower physical activity is associated with higher hippocampal CBF. For WM fMRI activation in the hippocampus, we observed a weak negative correlation with diet score (*r* = -0.17), indicating that better diet scores were associated with lower WM task-related brain activation in the hippocampus. For the variables of the cognitive functioning domain, we found that healthy diet score positively correlated with TMT performance (*r* = 0.15) and negatively with VFT performance (*r* = - 0.18). Lower LDL-cholesterol levels correlated with higher executive function measured by VFT (*r* = 0.16). A small correlation between WM fMRI activation in dlPFC and executive function measured by TMT was found (*r* = 0.16). Higher WM fMRI neural recruitment in the hippocampus was positively associated with episodic memory performance in the RAVLT delayed recall measure (*r* = 0.16).

**FIGURE 3.**
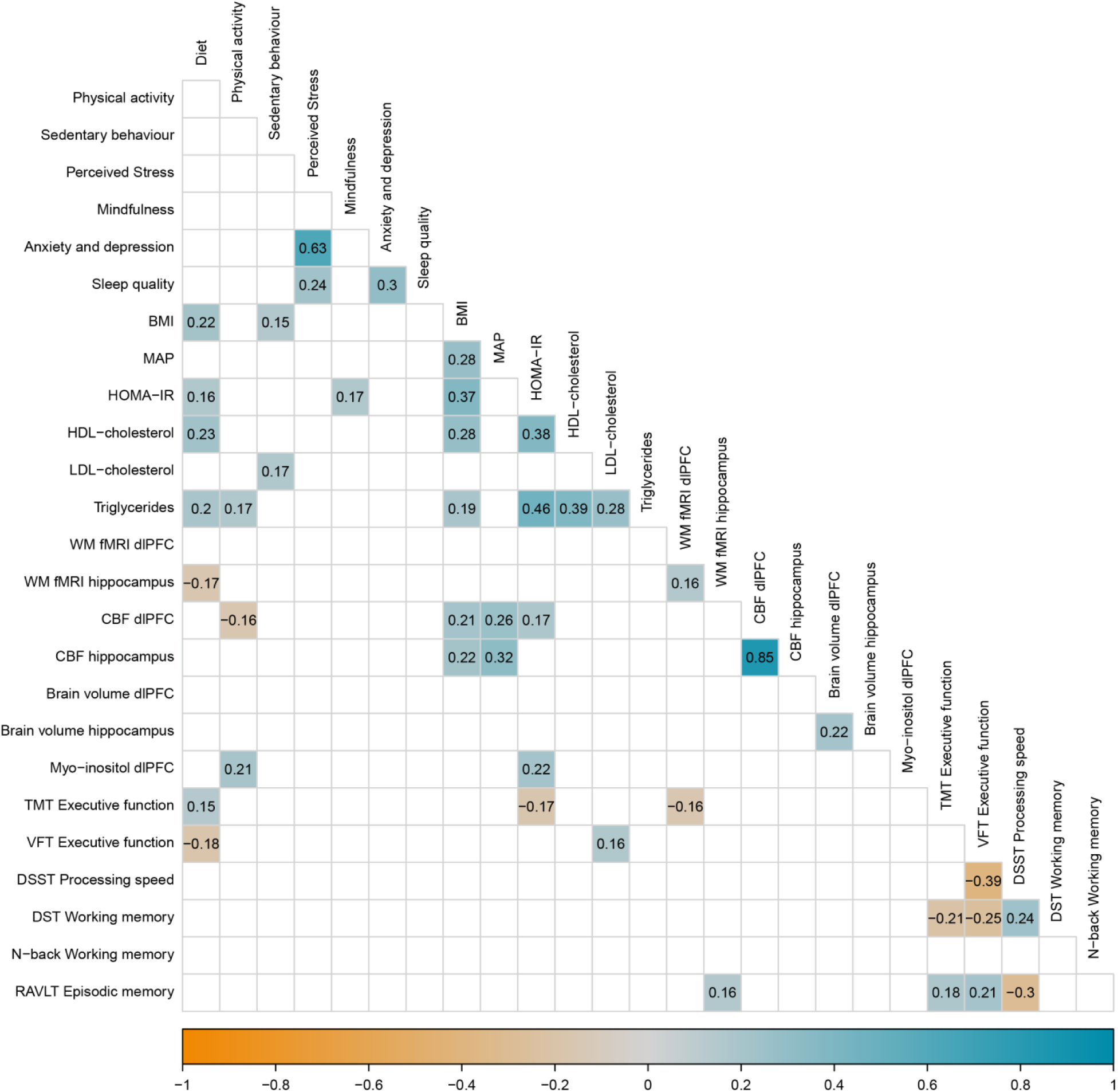
Pearson’s pairwise correlation matrix of all included variables. Only significant correlations (p <0.05) are shown. All variables have been transformed so a higher score indicates a better/healthier outcome. *Abbreviations:* BMI = body mass index; CBF = cerebral blood flow; DHD = Dutch Healthy Diet; dlPFC = dorsolateral prefrontal cortex; DSST = Digit Symbol Substitution Test; DST = Digit Span Test; FFMQ = Five Faceted Mindfulness Questionnaire; fMRI = functional magnetic resonance imaging; HADS = Hospital and Anxiety Depression Scale; HDL = high-density lipoproteins; HOMA-IR = Homeostatic Model Assessment for Insulin Resistance; LDL = low-density lipoproteins; MAP = mean arterial pressure; PSQI = Pittsburgh sleep quality index; PSS = Perceived Stress Scale; SBQ = Sedentary Behavior Questionnaire; SQUASH = Short Questionnaire to Assess Health-enhancing physical activity; RAVLT = Rey Auditory Verbal Learning Test; TMT = Trail Making Test; VFT = Verbal Fluency Test; WM = working memory.

### 3.6 Exploratory analyses for males and females

Because of the observed significant effects of sex (female) as a covariate in SEM model with the strongest model fit, we performed post-hoc analyses in subsets to further explore differences in associations between the total domain scores, as well as the individual outcome measures between men and women. In women, simple linear regression models showed a significant association between the composite domains of lifestyle and vascular health (*β* = 0.55, *SE* = 0.17, *p* = 0.002), and a trend-significant association between the composite domains of vascular health and brain health (*β* = 0.15, *SE* = 0.08, *p* = 0.058), as opposed to no significant correlations in men (*p* > 0.34) (Appendix A: Supplementary Figure 1). In the pairwise Pearson’s correlation analyses, women showed more positive associations between diet score and vascular health measures of BMI (*r* = 0.27), HOMA-IR (*r* = 0.26) and triglycerides (*r* = 0.21) compared to men. Compared to women, men showed more positive associations between vascular health measures BMI and HOMA-IR in measures of CBF within the dLPFC (*r* = 0.30 – 0.31) and hippocampus (*r* = 0.33). The complete results of the total domain simple linear regression models, as well as the individual outcome measure Pearson’s correlation analysis, are presented in Appendix A: Supplementary Figures 1-3.

## 4. DISCUSSION

In this cross-sectional exploratory study in older adults, we investigated a model integrating multiple facets of lifestyle, vascular health, brain health and cognitive functioning to identify possible correlational pathways and to isolate the strongest variables driving the shared variance between these domains. We hypothesized that we would find a pathway in a sequential order of the domain of lifestyle correlating with vascular health, which in turn correlates with brain health, which ultimately is associated with cognitive functioning. We found significant weak correlations between the lifestyle and vascular health domain scores, and between vascular health and brain, but observed no significant association between the domains of brain health and cognitive functioning. Except for the latter non-significant brain-cognition association, these findings are in line with our primary hypothesis of sequential effects. The integration of any additional direct pathways between the domains (e.g., from vascular health to cognitive functioning) did not improve the model. We furthermore found that the CCA models confirmed the established link between vascular and brain health, with CBF in the dlPFC and hippocampus emerging as the strongest contributors to shared variance within this population. These results were replicated in bivariate correlations between individual outcomes of vascular and brain health.

Our observed sequential links between lifestyle patterns, vascular health, brain health, and, subsequently, cognitive health, are largely in line with previous studies that have found bivariate links between lifestyle and vascular health, vascular health and brain health, or brain health and cognitive functioning in aging [3, 37–41]. Although we were unable to find a statistically significant association between the collective set of brain health markers and cognitive functioning, the observed coefficients did point in our hypothesized direction (i.e. healthier outcomes of brain structure and function are collectively associated with higher cognitive functioning). This final step, the translation from neuroimaging brain health markers to cognitive functioning measured by neuropsychological tests, is notoriously difficult [42]. Nevertheless, multiple studies have reported associations between measures of brain health and cognitive performance in aging, such as increased fMRI responses [43], CBF [11, 44], global and local brain volumes [9, 45, 46], and neuroinflammation [47]. Our observed inconsistent findings compared to previous studies may be explained by differences in study populations, as these prior studies generally included populations significantly older [9, 44, 45] or younger than 60-75 years [43, 46], or individuals with diagnosed mild cognitive impairment or Alzheimer’s Disease [11, 47]. Additionally, many of these studies consisted of longitudinal data or much larger sample sizes which, compared to the cross-sectional design and limited sample size of the current study, may have limited our ability to reduce between-individual variability and detect subtle associations.

Although direct associations between healthy lifestyle patterns on brain health markers or cognitive functioning have been reported previously [48, 49], we did not observe similar associations. Lifestyle markers showed minimal association with markers of brain health or cognitive functioning, both when examined individually and when combined into a composite domain score, except for mixed associations between dietary score and measures of executive functioning performance indicating task-specific effects on executive function performance. Additionally, our SEM model analysis indicated that the addition of any direct pathways between lifestyle and brain health, lifestyle and cognitive functioning, or vascular health and cognitive functioning, directly lowered the model’s fit. In our population of older at-risk adults, the impact of lifestyle on brain health (and potentially cognitive functioning) was strongest when mediated through vascular health. This finding is in line with previous studies, which indicate strong correlations between cardiovascular risk and specific brain regions involved in higher-order cognitive functions, such as the dlPFC and hippocampus, which support executive functioning, working memory, and episodic memory [6, 8]. The mixed findings between our analysis and previously reported protective factors of lifestyle on cognition, could be due to methodological differences. Our cross-sectional analysis approach is not able to capture long-term or causal effects, as opposed to longitudinal intervention studies [48, 50, 51]. However, not all longitudinal lifestyle intervention studies found direct effects of positive lifestyle changes on measures of cognitive functioning, which suggests that the relationship between lifestyle and cognition is still not fully understood [52–54] and might benefit from the inclusion of vascular factors as mediator.

The observed variance of multiple vascular health markers such as BMI, blood pressure (MAP), and blood-lipid markers with brain health markers was predominantly expressed in shared variance with CBF within the dlPFC and hippocampus. This relatively strong association between cardiovascular health and our CBF measures is in line with the sensitivity of the dlPFC and hippocampus structures to cardiovascular burden [6, 8, 55]; however, we did not include whole-brain CBF measures, which could display similar associations. We also found that participants with unhealthier scores of BMI, MAP, HOMA-IR, LDL-cholesterol and triglycerides had higher dlPFC volumes. This finding was unexpected, as the (dl)PFC has been reported to be negatively affected by such cardiovascular risk factors in an earlier longitudinal follow-up study in healthy and at-risk adults [56]. Although current literature does not yet offer an explanation for this finding, it is possible that our cross-sectional analysis was not able to capture the long-term detrimental effects of cardiovascular risk on dlPFC. This notion is supported by the lack of significant bivariate correlations between these vascular and volumetric outcomes, suggesting that volumetric outcomes may be less robust than the vascular-CBF links which were consistently significant across all our analyses.

We did not find any significant bivariate associations within the models relating lifestyle and vascular health markers, and brain health and cognitive functioning markers. The associations found when using total domain scores were largely lost within the CCA models where all variables of each set were individually weighted. This finding potentially points to an additive effect where the whole effect of all variables within the different sets (i.e., the z-score summed sets) was greater than the sum of its parts (i.e., the individual markers). Within the context of lifestyle effects on cognitive decline, previous studies, including a meta-analysis, have shown that lifestyle interventions targeting multiple different lifestyle domains have broader positive effects on cardiovascular risk factors than single-domain interventions [57, 58]. This suggests a potential additive effect: targeting multiple mechanisms underlying cognitive aging may improve overall intervention effectiveness. The latent composite domain score used in the current study, based on the sums of z-scaled markers, can provide a more robust and simplified way to compare multiple different outcome measures compared to single metrics and more closely follows the multifactorial etiology of aging-related cognitive decline and dementia. Our approach could especially be useful when grouping measures that assess the same underlying domain. Therefore, the present analysis performed on this latent domain variable could provide significant results, which were lost when looking at the individual components used to calculate the composite score.

Our study has several limitations. We performed exploratory data analyses with a cross-sectional approach. Therefore, even though our analyses revealed significant relations between a number of lifestyle, vascular health and brain health markers, the possibility of an unknown variable influencing these associations cannot be ruled out completely. Additionally, although evidence of the directionality of the hypothesized pathways and associations is robust, we are unable to formulate conclusions of possible causal relations between the observed associations. Another point of caution concerns our created brain health score, where we used higher dlPFC and hippocampal fMRI activation during the working memory N-back task as indicators of better brain health. Although the dlPFC and hippocampus are strongly associated with working memory-related tasks [59, 60], both brain regions show distinct task-related activation patterns which at times are non-parallel and may indicate different cognitive processes [61]. Therefore, one could argue that a higher neural recruitment in the dlPFC and hippocampus does not necessarily indicate a healthier brain outcome. Multiple studies have reported conflicting evidence of differences in hyper- or hypoactivation between healthy younger and older adults [62, 63], as well as in patients with mild cognitive impairment or dementia [64]. The possibility of compensatory mechanisms and individual differences influencing brain activation should be considered, especially considering the older age of our study population and their inclusion based on multiple cardiovascular risk factors. To add to this, we found one association which could possibly point to reduced compensatory need, where participants with lower WM fMRI activation in the hippocampus adhered to healthier dietary patterns. Another limitation of our study is that we only observed small associations between variables or domain composite scores. Therefore, interpretation of these results should be cautious and nuanced. One possible explanation for the lack of strong associations is the heterogeneity in risk profiles within our study sample. Because participants were at-risk of cognitive decline, comparisons between individuals with, for example, low versus high blood pressure do not necessarily represent a contrast between unhealthy and healthy individuals, as participants with lower blood pressure by definition had other risk factors. This lack of a healthy control group and the heterogeneity of individual risk factor profiles may have attenuated the observed associations. Lastly, it should be noted that we only analyzed simple linear relationships between individual markers and total domain scores, leading to possible oversimplified conclusions. With our current analysis, we were unable to examine potential non-linear, quadratic, or moderation effects among our included outcome variables. Thereby, more complex associations were not tested, and we only examined the possible direct and indirect pathways of our hypothesized sequential pathway order. As part of further exploratory analyses, other sequential orders (e.g., lifestyle-brain health associations mediated through cognitive functioning or vascular health markers) could be considered.

In conclusion, our study has extended previous studies by integrating multiple outcome measures of lifestyle, vascular health, brain health, and cognitive functioning into one integrated model to investigate the potential partial- or full mediation pathway of lifestyle ultimately affecting brain health and cognitive functioning. Our findings indicate a possible additive effect of multiple facets of lifestyle through vascular health on brain health. Future lifestyle intervention studies on such vascular, brain and cognitive markers, will offer more insight into their causal relationships in aging.

## Data Availability

Data available upon request.

## GLOSSARY

aCompCor: anatomical principal component noise regressors
AIC: Akaike Information Criterion
ASL: arterial spin labelling
BIC: Bayesian Information Criterion
BMI: body mass index
BOLD: blood-oxygen-level-dependent
CAIDE: Cardiovascular Risk Factors, Aging, and Incidence of Dementia
CBF: cerebral blood flow
CCA: canonical correlation analysis
CFI: Comparative Fit Index
CHESS: chemically selective water suppression
CSF: cerebrospinal fluid
DBP: diastolic blood pressure
DHD: Dutch Healthy Diet
dlPFC: dorsolateral prefrontal cortex
DSST: Digit Symbol Substitution Test
DST: Digit Span Test
EPI: echo planar imaging
FFMQ: Five Facet Mindfulness Questionnaire
FFQ: Food Frequency Questionnaire
fMRI: functional magnetic resonance imaging
FWHM: full width half maximum
GLM: general linear model
HADS: Hospital Anxiety and Depression Scale
HDL: high-density lipoprotein
HOMA-IR: Homeostatic Model Assessment for Insulin Resistance
ICV: intracranial volume
IQR: interquartile range
ISCED: International Standard Classification of Education
LDL: low-density lipoprotein
MAP: mean arterial pressure
MET: metabolic equivalent of task
MIND-NL: Dutch Mediterranean-Dietary Approaches to Stop Hypertension Intervention for Neurodegenerative Delay
pCASL: pseudocontinious arterial spin labelling
PRESS: Point RESolved Spectroscopy
PSQI: Pittsburgh Sleep Quality Index
PSS: Perceived Stress Scale
RAVLT: Rey Auditory Verbal Learning Test relResA relative residual amplitude
RMSEA: Root Mean Square Error of Approximation
ROIs: regions of interest
RU-DCCN: Radboud University Donders Center for Cognitive Neuroimaging
SBP: systolic blood pressure
LASA-SBQ: LASA Sedentary Behavior Questionnaire
SD: standard deviations
SEM: structural equation modelling
SNR: signal-to-noise ratio
SQUASH: Short Questionnaire to Assess Health-enhancing physical activity
tCompCor: temporal principal component noise regressors
TICS-M1: Telephone Interview for Cognitive Status (Modified version)
TMT: Trail Making Test
VFT: Verbal Fluency Test
WHO: World Health Organization
WM: working memory
WUR-HNH: Wageningen University department of Human Nutrition and Health

## ACKNOWLEDGEMENTS

We wish to thank Jill Naaijen, Viola Hollestein, and Nick Puts for their help with setting up the MRS MRI-sequence during the design phase of our study, and helping with the pre-processing, analysis and interpretation of the MRS data. We thank Nils Kohn for his valuable input on the statistical methodology used in this study. We thank Noortje Overwater for her help in coordinating data collection at the WUR-HNH. We thank all bachelor’s and master’s students who have helped with data collection. We also wish to express our sincerest thanks to all participants for their valuable time and commitment to this study.

This work was supported by a Crossover grant (MOCIA 17611) of the Dutch Research Council (NWO), granted in December 2019. The MOCIA program is a public-private partnership [12].

## AUTHOR CONTRIBUTIONS

**Mark R. van Loenen**: Conceptualization, Methodology, Validation, Formal Analysis, Investigation, Data Curation, Writing – Original Draft, Writing – Review & Editing, Visualization, Project Administration. **Lianne B. Remie**: Validation, Investigation, Data Curation, Writing – Original Draft, , Writing – Review & Editing, Project Administration. **Mara P.H. van Trijp**: Investigation, Data Curation, Writing – Review & Editing, Project Administration. **Mechteld M. Grootte Bromhaar**: Investigation, Data Curation, Project Administration. **José P. Marques**: Resources, Data Curation, Writing – Review & Editing, Funding Acquisition. **Jurgen A.H.R. Claassen**: Writing – Review & Editing, Funding Acquisition. **Wilma T. Steegenga**: Conceptualization, Methodology, Supervision, Project Administration, Funding Acquisition. **Joukje M. Oosterman**: Conceptualization, Methodology, Validation, Writing – Review & Editing, Supervision, Project Administration, Funding Acquisition. **Esther Aarts**: Conceptualization, Methodology, Validation, Writing – Review & Editing, Supervision, Project Administration, Funding Acquisition.

## REFERENCES

1. Organization, G.W.H., Risk reduction of cognitive decline and dementia: WHO guidelines. 2019: https://www.ncbi.nlm.nih.gov/books/NBK542796/. p. 96.

2. Kivipelto, M., F. Mangialasche, and T. Ngandu, Lifestyle interventions to prevent cognitive impairment, dementia and Alzheimer disease. Nature Reviews Neurology, 2018. 14(11): p. 653–666.

3. Livingston, G., et al., *Dementia prevention, intervention, and care:* 2024 report of the Lancet standing Commission. The Lancet, 2024. 404(10452): p. 572–628.

4. Sisti, L.G., et al., The effect of multifactorial lifestyle interventions on cardiovascular risk factors: a systematic review and meta-analysis of trials conducted in the general population and high risk groups. Prev Med, 2018. 109: p. 82–97.

5. Gorelick, P.B., et al., Vascular contributions to cognitive impairment and dementia: a statement for healthcare professionals from the american heart association/american stroke association. Stroke, 2011. 42(9): p. 2672–713.

6. Murman, D.L., The Impact of Age on Cognition. Semin Hear, 2015. 36(3): p. 111–21.

7. Launer, L.J., et al., Vascular Factors and Multiple Measures of Early Brain Health: CARDIA Brain MRI Study. PLOS ONE, 2015. 10(3): p. e0122138.

8. Fjell, A.M., et al., What is normal in normal aging? Effects of aging, amyloid and Alzheimer’s disease on the cerebral cortex and the hippocampus. Prog Neurobiol, 2014. 117: p. 20–40.

9. Cardenas, V.A., et al., Brain atrophy associated with baseline and longitudinal measures of cognition. Neurobiol Aging, 2011. 32(4): p. 572–80.

10. Gorbach, T., et al., Longitudinal association between hippocampus atrophy and episodic-memory decline. Neurobiol Aging, 2017. 51: p. 167–176.

11. De Vis, J.B., et al., Arterial-spin-labeling (ASL) perfusion MRI predicts cognitive function in elderly individuals: A 4-year longitudinal study. J Magn Reson Imaging, 2018. 48(2): p. 449–458.

12. MOCIA. *MOCIA Scientific*. 2024 [cited 2024; Available from: https://mocia.nl/scientific/.

13. van Loenen, M.R., et al., The effects of a multidomain lifestyle intervention on brain functioning and its relation with immunometabolic markers and intestinal health in older adults at-risk of cognitive decline: study design and baseline characteristics of the HELI randomized controlled trial. JMIR Research Protocols, 2025(69814 (forthcoming/in press)).

14. Kivipelto, M., et al., Risk score for the prediction of dementia risk in 20 years among middle aged people: a longitudinal, population-based study. Lancet Neurol, 2006. 5(9): p. 735–41.

15. van den Berg, E., et al., The Telephone Interview for Cognitive Status (Modified): relation with a comprehensive neuropsychological assessment. J Clin Exp Neuropsychol, 2012. 34(6): p. 598–605.

16. OECD, E.U., UNESCO Institute for Statistics, ISCED 2011: Operational Manual: Guidelines for Classifying National Education Programmes and Related Qualifications. 2011, OECD Publishing: 10.1787/9789264228368-en.

17. Kromhout, D., et al., The 2015 Dutch food-based dietary guidelines. Eur J Clin Nutr, 2016. 70(8): p. 869–78.

18. de Rijk, M.G., et al., Development and evaluation of a diet quality screener to assess adherence to the Dutch food-based dietary guidelines. Br J Nutr, 2021. 128(8): p. 1–11.

19. Beers, S., et al., Development of the Dutch Mediterranean-Dietary Approaches to Stop Hypertension Intervention for Neurodegenerative Delay (MIND) Diet and its scoring system, alongside the modification of a brief FFQ for assessing dietary adherence. Br J Nutr, 2024: p. 1–9.

20. Morris, M.C., et al., MIND diet slows cognitive decline with aging. Alzheimers Dement, 2015. 11(9): p. 1015–22.

21. van Lee, L., et al., The Dutch Healthy Diet index (DHD-index): an instrument to measure adherence to the Dutch Guidelines for a Healthy Diet. Nutrition Journal, 2012. 11(1): p. 49.

22. Visser, M. and A. Koster, Development of a questionnaire to assess sedentary time in older persons – a comparative study using accelerometry. BMC Geriatrics, 2013. 13(1): p. 80.

23. Cohen, S., T. Kamarck, and R. Mermelstein, A Global Measure of Perceived Stress. Journal of Health and Social Behavior, 1983. 24(4): p. 385–396.

24. Zigmond, A.S. and R.P. Snaith, The hospital anxiety and depression scale. Acta Psychiatr Scand, 1983. 67(6): p. 361–70.

25. Buysse, D.J., et al., The Pittsburgh Sleep Quality Index: a new instrument for psychiatric practice and research. Psychiatry Res, 1989. 28(2): p. 193–213.

26. Nath Kundu, R., S. Biswas, and M. Das, Mean Arterial Pressure Classification: A Better Tool for Statistical Interpretation of Blood Pressure Related Risk Covariates. Cardiology and Angiology: An International Journal, 2017. 6(1): p. 1–7.

27. Fujimoto, K., et al., Quantitative comparison of cortical surface reconstructions from MP2RAGE and multi-echo MPRAGE data at 3 and 7 T. Neuroimage, 2014. 90: p. 60–73.

28. Owen, A.M., et al., N-back working memory paradigm: a meta-analysis of normative functional neuroimaging studies. Hum Brain Mapp, 2005. 25(1): p. 46–59.

29. Esteban, O., et al., fMRIPrep: a robust preprocessing pipeline for functional MRI. Nat Methods, 2019. 16(1): p. 111–116.

30. Wang, H., et al., A coordinate-based meta-analysis of the n-back working memory paradigm using activation likelihood estimation. Brain and Cognition, 2019. 132: p. 1–12.

31. Jenkinson, M., et al., FSL. Neuroimage, 2012. 62(2): p. 782–90.

32. Chappell, M.A., et al., Variational Bayesian inference for a nonlinear forward model. Trans. Sig. Proc., 2009. 57(1): p. 223–236.

33. Chang, L., et al., Magnetic resonance spectroscopy to assess neuroinflammation and neuropathic pain. J Neuroimmune Pharmacol, 2013. 8(3): p. 576–93.

34. Siemens, H., Syngo MR B19, Basic Manual-Spectroscopy. 2012, Siemens Healthineers AG Munich, Germany.

35. Oeltzschner, G., et al., Osprey: Open-source processing, reconstruction & estimation of magnetic resonance spectroscopy data. Journal of Neuroscience Methods, 2020. 343: p. 108827.

36. Klose, U., In vivo proton spectroscopy in presence of eddy currents. Magn Reson Med, 1990. 14(1): p. 26–30.

37. Ghodeshwar, G.K., A. Dube, and D. Khobragade, Impact of Lifestyle Modifications on Cardiovascular Health: A Narrative Review. Cureus, 2023. 15(7): p. e42616.

38. Bliss, E.S., et al., Benefits of exercise training on cerebrovascular and cognitive function in ageing. Journal of Cerebral Blood Flow & Metabolism, 2020. 41(3): p. 447–470.

39. Iadecola, C. and R.F. Gottesman, Neurovascular and Cognitive Dysfunction in Hypertension. Circ Res, 2019. 124(7): p. 1025–1044.

40. Lockhart, S.N., et al., Cardiometabolic disorders are associated with reduced cerebral perfusion and white matter microstructure. Alzheimer’s & Dementia, 2021. 17(S1): p. e055791.

41. Barnes, J.N. and A.T. Corkery, Exercise Improves Vascular Function, but does this Translate to the Brain? Brain Plast, 2018. 4(1): p. 65–79.

42. Litwińczuk, M.C., et al., Relating Cognition to both Brain Structure and Function: A Systematic Review of Methods. Brain Connect, 2023. 13(3): p. 120–132.

43. Yaple, Z.A., W.D. Stevens, and M. Arsalidou, Meta-analyses of the n-back working memory task: fMRI evidence of age-related changes in prefrontal cortex involvement across the adult lifespan. Neuroimage, 2019. 196: p. 16–31.

44. Bangen, K.J., et al., Interactive effects of vascular risk burden and advanced age on cerebral blood flow. Front Aging Neurosci, 2014. 6: p. 159.

45. Hedden, T., et al., Multiple Brain Markers are Linked to Age-Related Variation in Cognition. Cereb Cortex, 2016. 26(4): p. 1388–400.

46. Vibha, D., et al., Brain Volumes and Longitudinal Cognitive Change: A Population-based Study. Alzheimer Dis Assoc Disord, 2018. 32(1): p. 43–49.

47. Lecca, D., et al., Role of chronic neuroinflammation in neuroplasticity and cognitive function: A hypothesis. Alzheimers Dement, 2022. 18(11): p. 2327–2340.

48. Kimura, N., et al., Lifestyle factors that affect cognitive function-a longitudinal objective analysis. Front Public Health, 2023. 11: p. 1215419.

49. Ngandu, T., et al., A 2 year multidomain intervention of diet, exercise, cognitive training, and vascular risk monitoring versus control to prevent cognitive decline in at-risk elderly people (FINGER): a randomised controlled trial. The Lancet, 2015. 385(9984): p. 2255–2263.

50. Ngandu, T., et al., A 2 year multidomain intervention of diet, exercise, cognitive training, and vascular risk monitoring versus control to prevent cognitive decline in at-risk elderly people (FINGER): a randomised controlled trial. Lancet, 2015. 385(9984): p. 2255–63.

51. Fu, M., et al., Identifying common disease trajectories of Alzheimer&#x2019;s disease with electronic health records. eBioMedicine, 2025. 118.

52. Sugimoto, T., et al., Multidomain Intervention Trial for Preventing Cognitive Decline among Older Adults with Type 2 Diabetes: J-MIND-Diabetes. J Prev Alzheimers Dis, 2024. 11(6): p. 1604–1614.

53. Andrieu, S., et al., Effect of long-term omega 3 polyunsaturated fatty acid supplementation with or without multidomain intervention on cognitive function in elderly adults with memory complaints (MAPT): a randomised, placebo-controlled trial. Lancet Neurol, 2017. 16(5): p. 377–389.

54. Zülke, A.E., et al., A multidomain intervention against cognitive decline in an at-risk-population in Germany: Results from the cluster-randomized AgeWell.de trial. Alzheimers Dement, 2024. 20(1): p. 615–628.

55. Salat, D.H., J.A. Kaye, and J.S. Janowsky, Prefrontal gray and white matter volumes in healthy aging and Alzheimer disease. Arch Neurol, 1999. 56(3): p. 338–44.

56. Raz, N., et al., Vascular health and longitudinal changes in brain and cognition in middle-aged and older adults. Neuropsychology, 2007. 21(2): p. 149–57.

57. Wesselman, L.M., et al., Web-Based Multidomain Lifestyle Programs for Brain Health: Comprehensive Overview and Meta-Analysis. JMIR Ment Health, 2019. 6(4): p. e12104.

58. Toman, J., B. Klímová, and M. Vališ, Multidomain Lifestyle Intervention Strategies for the Delay of Cognitive Impairment in Healthy Aging. Nutrients, 2018. 10(10).

59. Hoshi, Y., et al., Spatiotemporal characteristics of hemodynamic changes in the human lateral prefrontal cortex during working memory tasks. Neuroimage, 2003. 20(3): p. 1493–504.

60. Churchwell, J.C. and R.P. Kesner, Hippocampal-prefrontal dynamics in spatial working memory: interactions and independent parallel processing. Behav Brain Res, 2011. 225(2): p. 389–95.

61. Lee, I. and R.P. Kesner, Time-dependent relationship between the dorsal hippocampus and the prefrontal cortex in spatial memory. J Neurosci, 2003. 23(4): p. 1517–23.

62. Saliasi, E., et al., Neural correlates associated with successful working memory performance in older adults as revealed by spatial ICA. PLoS One, 2014. 9(6): p. e99250.

63. Park, D.C. and I.M. McDonough, The Dynamic Aging Mind: Revelations From Functional Neuroimaging Research. Perspectives on Psychological Science, 2013. 8(1): p. 62–67.

64. Terry, D.P., et al., A Meta-Analysis of fMRI Activation Differences during Episodic Memory in Alzheimer’s Disease and Mild Cognitive Impairment. J Neuroimaging, 2015. 25(6): p. 849–60.

