## Appendix: Supplementary Materials for "Integrating lifestyle, vascular, brain and cognitive markers: an exploratory multimodal approach to brain aging"

### Appendix A. Supplementary Materials

| **SUPPLEMENTARY TABLE 1A.** Tests of canonical dimensions of CCA model of lifestyle and vascular health domains. | | | | | | |
| --- | --- | --- | --- | --- | --- | --- |
| **Dimension** | **Rc** | **Rc^2^** | **Wilks’ λ** | ***F*-statistic** | **df** | ***p­*-value** |
| 1 | 0.353 | 0.125 | 0.739 | 1.011 | 42, 636.658 | 0.454 |
| 2 | 0.277 | 0.077 | 0.844 | 0.788 | 30, 546 | 0.784 |
| 3 | 0.207 | 0.043 | 0.914 | 0.623 | 20, 455.327 | 0.897 |
| 4 | 0.177 | 0.031 | 0.955 | 0.532 | 12, 365.405 | 0.894 |
| 5 | 0.113 | 0.013 | 0.986 | 0.323 | 6, 278 | 0.925 |
| 6 | 0.031 | 0.001 | 0.999 | 0.066 | 2, 140 | 0.936 |
| Canonical dimensions 1–6: Linear combinations of variables (dimensions) that maximize shared variance between two variable sets. Dimension 1 captures the strongest shared variance, followed by dimension 2, and so on. Canonical correlations (Rc), squared canonical correlations (Rc^2^), Wilks’ λ, F-statistic, degrees of freedom (df), and p-values for each canonical dimension between lifestyle and vascular health domains. | | | | | | |

| **SUPPLEMENTARY TABLE 1B.** Canonical loadings of all dimensions of CCA model of lifestyle and vascular health domains. | | | | | | |
| --- | --- | --- | --- | --- | --- | --- |
| **Variable** | **Dimension 1** | **Dimension 2** | **Dimension 3** | **Dimension 4** | **Dimension 5** | **Dimension 6** |
| **Set *X* (Lifestyle)** |  |  |  |  |  |  |
| DHD | -0.598 | 0.064 | -0.330 | 0.339 | -0.294 | 0.570 |
| SQUASH | -0.487 | -0.259 | -0.318 | -0.687 | -0.064 | -0.300 |
| LASA | -0.475 | 0.709 | 0.390 | -0.294 | 0.103 | -0.022 |
| PSS | -0.003 | 0.448 | -0.133 | 0.203 | -0.355 | -0.605 |
| HADS | -0.078 | 0.095 | 0.161 | 0.197 | 0.055 | -0.187 |
| FFMQ | -0.471 | -0.388 | 0.232 | 0.405 | 0.281 | -0.446 |
| PSQI | 0.159 | 0.279 | -0.630 | 0.187 | 0.605 | -0.090 |
| **Set *Y* (Vascular health)** |  |  |  |  |  |  |
| BMI | -0.770 | 0.053 | 0.383 | -0.192 | -0.469 | 0.032 |
| MAP | -0.166 | 0.098 | 0.472 | -0.686 | 0.349 | 0.383 |
| HOMA-IR | -0.695 | -0.530 | -0.116 | 0.229 | 0.189 | 0.366 |
| HDL-cholesterol | -0.459 | 0.315 | -0.582 | -0.001 | -0.251 | 0.536 |
| LDL-cholesterol | -0.260 | 0.633 | 0.285 | 0.362 | 0.538 | -0.171 |
| Triglycerides | -0.706 | 0.073 | -0.453 | -0.208 | 0.455 | -0.200 |
| Canonical loadings (correlation between each variable and the canonical variate) of each canonical dimension for the domains of lifestyle (set *X*) and vascular health (set *Y*). Data was Z-scored, and select variables were transformed so higher Z-scores indicate healthier outcomes.  *Abbreviations:* BMI = body mass index; DHD = Dutch Healthy Diet; FFMQ = Five Faceted Mindfulness Questionnaire; HADS = Hospital and Anxiety Depression Scale; HDL = high-density lipoproteins; HOMA-IR = Homeostatic Model Assessment for Insulin Resistance; LDL = low-density lipoproteins; MAP = mean arterial pressure; PSQI = Pittsburgh sleep quality index; PSS = Perceived Stress Scale; SBQ = Sedentary Behavior Questionnaire; SQUASH = Short Questionnaire to Assess Health-enhancing physical activity. | | | | | | |

| **SUPPLEMENTARY TABLE 2A.** Tests of canonical dimensions of CCA model of vascular health and brain health domains. | | | | | | |
| --- | --- | --- | --- | --- | --- | --- |
| **Dimension** | **Rc** | **Rc^2^** | **Wilks’ λ** | ***F*-statistic** | **df** | ***p­*-value** |
| 1 | 0.431 | 0.186 | 0.633 | 1.336 | 42, 547.540 | 0.081 |
| 2 | 0.324 | 0.105 | 0.777 | 1.019 | 30, 470 | 0.441 |
| 3 | 0.273 | 0.075 | 0.868 | 0.854 | 20, 392.312 | 0.646 |
| 4 | 0.195 | 0.038 | 0.938 | 0.640 | 12, 315.136 | 0.807 |
| 5 | 0.154 | 0.024 | 0.975 | 0.501 | 6, 240 | 0.807 |
| 6 | 0.032 | 0.001 | 0.999 | 0.061 | 2, 121 | 0.941 |
| Canonical dimensions 1–6: Linear combinations of variables (dimensions) that maximize shared variance between two variable sets. Dimension 1 captures the strongest shared variance, followed by dimension 2, and so on. Canonical correlations (Rc), squared canonical correlations (Rc^2^), Wilks’ λ, F-statistic, degrees of freedom (df), and p-values for each canonical dimension between vascular health and brain health domains. | | | | | | |

| **SUPPLEMENTARY TABLE 2B.** Canonical loadings of all dimensions of CCA model of vascular health and brain health domains. | | | | | | |
| --- | --- | --- | --- | --- | --- | --- |
| **Variable** | **Dimension 1** | **Dimension 2** | **Dimension 3** | **Dimension 4** | **Dimension 5** | **Dimension 6** |
| **Set *X* (Vascular health)** |  |  |  |  |  |  |
| BMI | -0.370 | 0.659 | 0.255 | 0.477 | -0.149 | 0.336 |
| MAP | -0.837 | -0.204 | 0.007 | 0.308 | -0.319 | -0.247 |
| HOMA-IR | -0.373 | 0.612 | 0.123 | 0.055 | 0.653 | -0.206 |
| HDL-cholesterol | 0.108 | -0.012 | 0.864 | 0.355 | 0.338 | -0.041 |
| LDL-cholesterol | -0.408 | -0.380 | -0.102 | -0.070 | 0.345 | 0.745 |
| Triglycerides | -0.466 | 0.125 | 0.653 | -0.522 | 0.193 | 0.178 |
| **Set *Y* (Brain health)** |  |  |  |  |  |  |
| WM fMRI activation dlPFC | 0.065 | 0.231 | -0.270 | 0.231 | -0.425 | -0.118 |
| WM fMRI activation hippocampus | -0.101 | -0.266 | -0.424 | -0.076 | 0.109 | 0.743 |
| CBF dlPFC | -0.050 | 0.821 | 0.066 | -0.424 | 0.250 | 0.274 |
| CBF hippocampus | -0.738 | 0.010 | 0.142 | 0.518 | 0.390 | 0.101 |
| Volume dlPFC | -0.844 | 0.039 | -0.086 | 0.420 | -0.011 | 0.195 |
| Volume hippocampus | 0.499 | 0.111 | -0.080 | 0.740 | 0.023 | 0.262 |
| Myo-inositol dlPFC | 0.064 | 0.000 | 0.777 | 0.175 | -0.332 | 0.488 |
| Canonical loadings (correlation between each variable and the canonical variate) of each canonical dimension for the domains of vascular health (set *X*) and brain health (set *Y*). Data was Z-scored, and select variables were transformed so higher Z-scores indicate healthier outcomes. Positive canonical loadings correspond to healthier scores, whereas negative canonical loadings correspond to less healthy scores.  *Abbreviations:* BMI = body mass index; CBF = cerebral blood flow; dlPFC = dorsolateral prefrontal cortex; fMRI = functional magnetic resonance imaging; HADS = Hospital and Anxiety Depression Scale; HDL = high-density lipoproteins; HOMA-IR = Homeostatic Model Assessment for Insulin Resistance; LDL = low-density lipoproteins; MAP = mean arterial pressure; WM = working memory. | | | | | | |

| **SUPPLEMENTARY TABLE 3A.** Tests of canonical dimensions of CCA model of brain health and cognitive functioning domains. | | | | | | |
| --- | --- | --- | --- | --- | --- | --- |
| **Dimension** | **Rc** | **Rc^2^** | **Wilks’ λ** | ***F*-statistic** | **df** | ***p­*-value** |
| 1 | 0.305 | 0.093 | 0.795 | 0.730 | 42, 613.206 | 0.897 |
| 2 | 0.260 | 0.068 | 0.877 | 0.585 | 30, 526 | 0.963 |
| 3 | 0.201 | 0.040 | 0.940 | 0.410 | 20, 438.744 | 0.990 |
| 4 | 0.120 | 0.014 | 0.980 | 0.224 | 12, 352.176 | 0.997 |
| 5 | 0.074 | 0.005 | 0.994 | 0.127 | 6, 268 | 0.993 |
| 6 | 0.014 | 0.000 | 1.000 | 0.012 | 2, 135 | 0.988 |
| Canonical dimensions 1–6: Linear combinations of variables (dimensions) that maximize shared variance between two variable sets. Dimension 1 captures the strongest shared variance, followed by dimension 2, and so on. Canonical correlations (Rc), squared canonical correlations (Rc^2^), Wilks’ λ, F-statistic, degrees of freedom (df), and p-values for each canonical dimension between brain health and cognitive functioning domains. | | | | | | |

| **SUPPLEMENTARY TABLE 3B.** Canonical loadings of all dimensions of CCA model of brain health and cognitive functioning domains. | | | | | | |
| --- | --- | --- | --- | --- | --- | --- |
| Variable | **Dimension 1** | **Dimension 2** | **Dimension 3** | **Dimension 4** | **Dimension 5** | **Dimension 6** |
| **Set *X* (Brain health)** |  |  |  |  |  |  |
| WM fMRI activation dlPFC | -0.360 | 0.522 | 0.217 | -0.459 | 0.263 | 0.439 |
| WM fMRI activation hippocampus | 0.310 | -0.193 | 0.711 | -0.352 | -0.253 | 0.415 |
| CBFdlLPFC | 0.396 | 0.568 | 0.416 | 0.316 | -0.172 | -0.467 |
| CBF hippocampus | -0.021 | 0.332 | 0.679 | 0.384 | -0.245 | -0.468 |
| Volume dlPFC | -0.055 | 0.004 | -0.140 | -0.664 | -0.329 | -0.649 |
| Volume hippocampus | 0.104 | -0.071 | -0.070 | 0.047 | -0.388 | -0.019 |
| Myo-inositol dlPFC | -0.111 | 0.234 | -0.307 | 0.244 | -0.709 | 0.438 |
| **Set *Y* (Cognitive functioning)** |  |  |  |  |  |  |
| TMT Executive function | 0.317 | -0.276 | -0.125 | 0.872 | -0.215 | -0.029 |
| VFT Executive function | 0.572 | -0.269 | -0.070 | -0.135 | 0.657 | -0.381 |
| DSST Processing speed | -0.575 | 0.650 | -0.081 | 0.444 | 0.102 | -0.179 |
| DST Working memory | 0.320 | 0.788 | 0.185 | -0.030 | -0.071 | 0.486 |
| N-back Working memory | -0.333 | 0.007 | -0.148 | 0.040 | 0.547 | 0.753 |
| RAVLT Episodic memory | 0.194 | -0.219 | 0.940 | 0.133 | 0.098 | -0.061 |
| Canonical loadings (correlation between each variable and the canonical variate) of each canonical dimension for the domains of brain health (set *X*) and cognitive functioning (set *Y*). Data was Z-scored, and select variables were transformed so higher Z-scores indicate healthier outcomes. Positive canonical loadings correspond to healthier scores, whereas negative canonical loadings correspond to less healthy scores.  *Abbreviations:* CBF = cerebral blood flow; dlPFC = dorsolateral prefrontal cortex; DSST = Digit Symbol Substitution Test; DST = Digit Span Test; fMRI = functional magnetic resonance imaging; RAVLT = Rey Auditory Verbal Learning Test; TMT = Trail Making Test; VFT = Verbal Fluency Test; WM = working memory. | | | | | | |

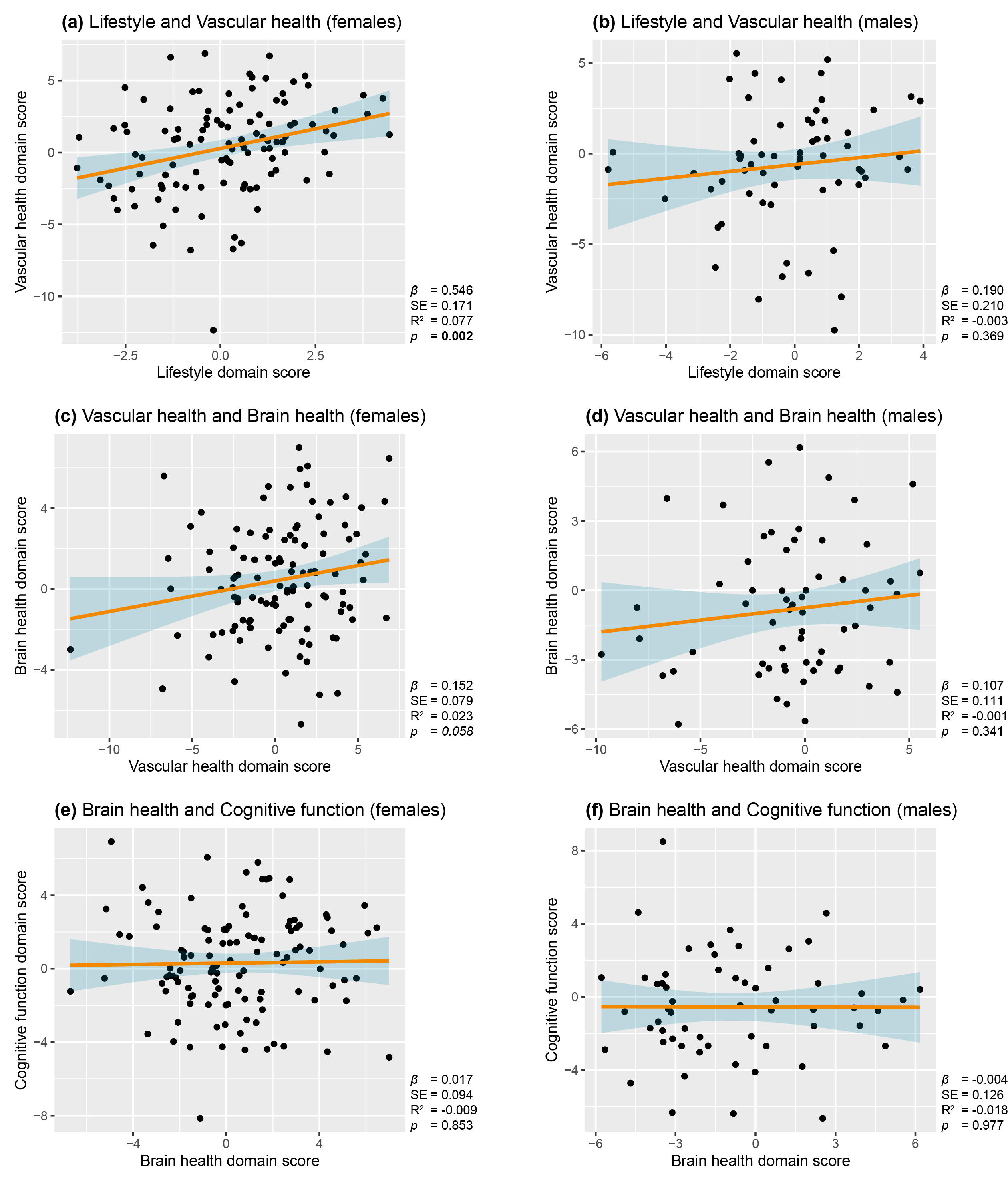

Supplementary Figure 1. Associations between z-summed total domain scores in Female vs. Male subsets.
Linear regression plots showing relationships between domain scores of lifestyle and vascular health, vascular health and brain health, and brain health and cognitive functioning in Female (panels a, c, and e) and Male (panels b, d, and f) subsets. The 95% confidence interval is indicated by shaded areas. R^2^ is reported as adjusted R^2^.

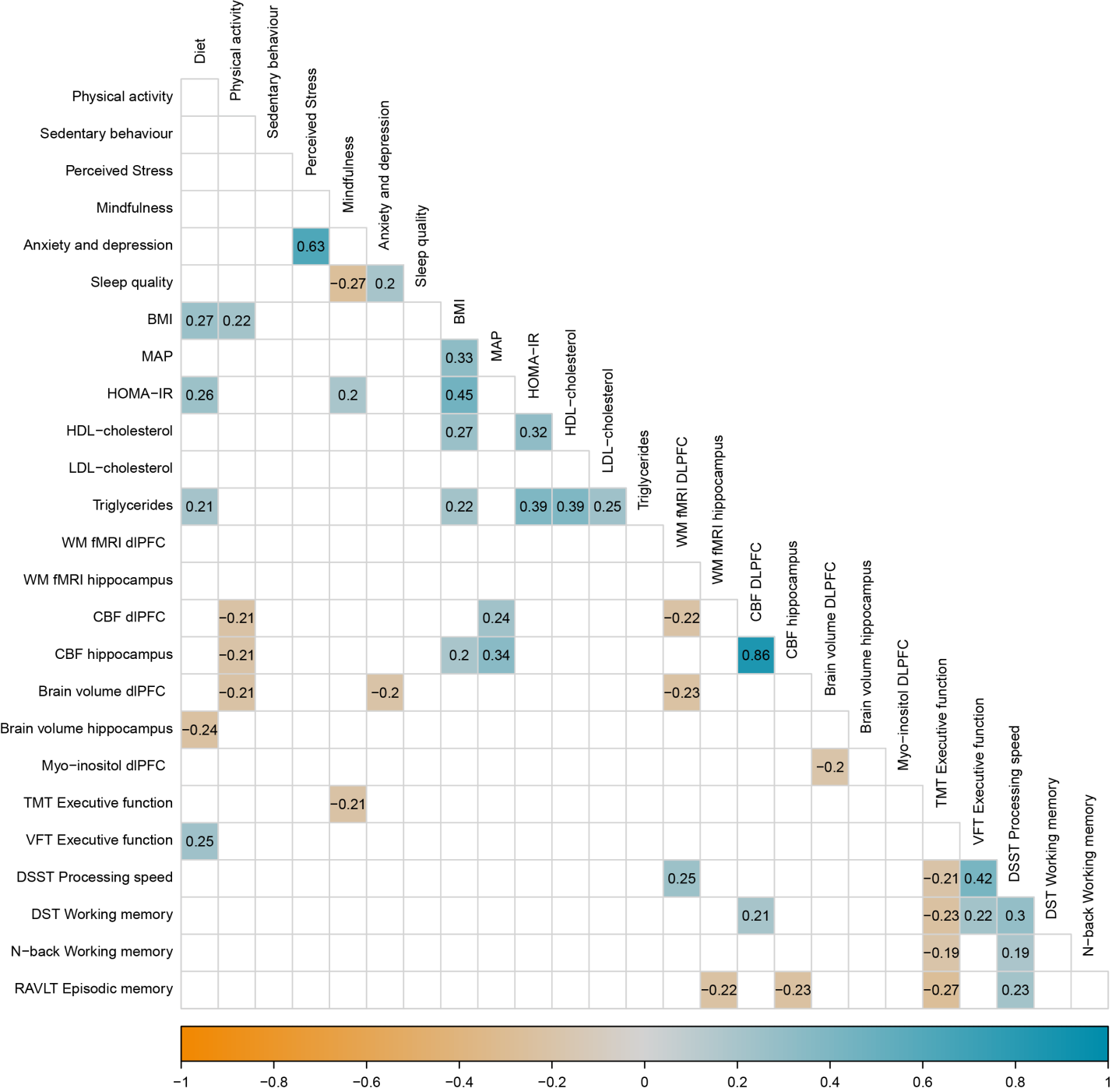

SUPPLEMENTARY FIGURE 2. Female subset correlation matrix of all included lifestyle, vascular, brain and cognitive functioning outcome measures. Only significant correlations (p <0.05) are shown. Select variables were transformed so higher Z-scores indicate healthier outcomes.

*Abbreviations:* BMI = body mass index; CBF = cerebral blood flow; DHD = Dutch Healthy Diet; dlPFC = dorsolateral prefrontal cortex; DSST = Digit Symbol Substitution Test; DST = Digit Span Test; FFMQ = Five Faceted Mindfulness Questionnaire; fMRI = functional magnetic resonance imaging; HADS = Hospital and Anxiety Depression Scale; HDL = high-density lipoproteins; HOMA-IR = Homeostatic Model Assessment for Insulin Resistance; LDL = low-density lipoproteins; MAP = mean arterial pressure; PSQI = Pittsburgh sleep quality index; PSS = Perceived Stress Scale; SBQ = Sedentary Behavior Questionnaire; SQUASH = Short Questionnaire to Assess Health-enhancing physical activity; RAVLT = Rey Auditory Verbal Learning Test; TMT = Trail Making Test; VFT = Verbal Fluency Test; WM = working memory.

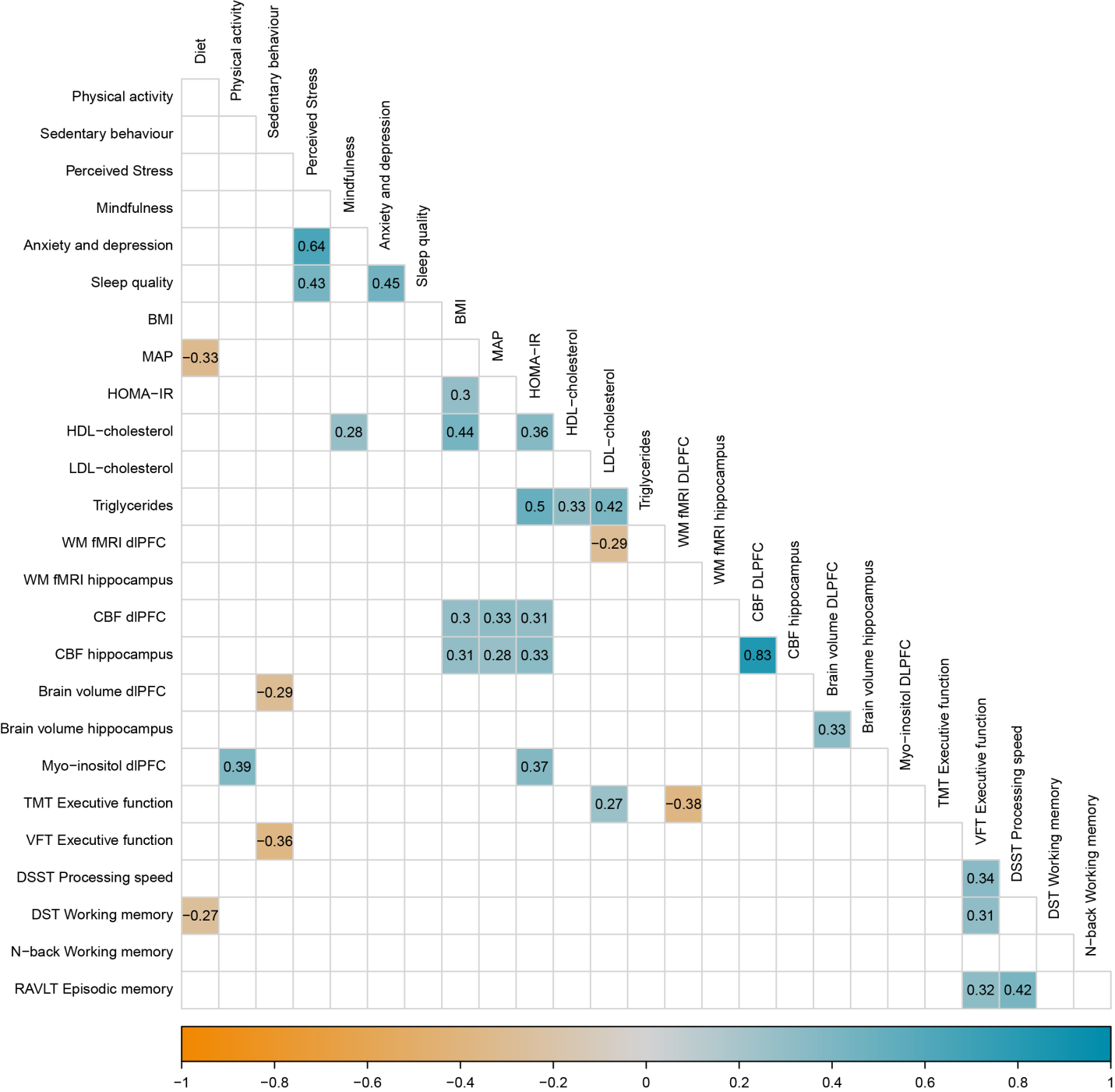

SUPPLEMENTARY FIGURE 3. Male subset correlation matrix of all included lifestyle, vascular, brain and cognitive functioning outcome measures. Only significant correlations (p <0.05) are shown. Select variables were transformed so higher Z-scores indicate healthier outcomes.

*Abbreviations:* BMI = body mass index; CBF = cerebral blood flow; DHD = Dutch Healthy Diet; dlPFC = dorsolateral prefrontal cortex; DSST = Digit Symbol Substitution Test; DST = Digit Span Test; FFMQ = Five Faceted Mindfulness Questionnaire; fMRI = functional magnetic resonance imaging; HADS = Hospital and Anxiety Depression Scale; HDL = high-density lipoproteins; HOMA-IR = Homeostatic Model Assessment for Insulin Resistance; LDL = low-density lipoproteins; MAP = mean arterial pressure; PSQI = Pittsburgh sleep quality index; PSS = Perceived Stress Scale; SBQ = Sedentary Behavior Questionnaire; SQUASH = Short Questionnaire to Assess Health-enhancing physical activity; RAVLT = Rey Auditory Verbal Learning Test; TMT = Trail Making Test; VFT = Verbal Fluency Test; WM = working memory.
